# Comparison of four methods to measure haemoglobin concentrations in whole blood donors (COMPARE): a diagnostic accuracy study

**DOI:** 10.1101/2020.11.06.20226779

**Authors:** Steven Bell, Michael Sweeting, Anna Ramond, Ryan Chung, Stephen Kaptoge, Matthew Walker, Thomas Bolton, Jennifer Sambrook, Carmel Moore, Amy McMahon, Sarah Fahle, Donna Cullen, Susan Mehenny, Angela M Wood, Jane Armitage, Willem H Ouwehand, Gail Miflin, Dave J Roberts, John Danesh, Emanuele Di Angelantonio, on behalf of the COMPARE Study Group

## Abstract

**Objective:** To compare four haemoglobin measurement methods in whole blood donors.

**Background:** To safeguard donors, blood services measure haemoglobin concentration in advance of each donation. NHS Blood and Transplant’s (NHSBT) usual method has been capillary gravimetry (copper sulphate), followed by venous HemoCue® (spectrophotometry) for donors failing gravimetry. However, gravimetry/venous HemoCue® results in 10% of donors being inappropriately bled (i.e., with haemoglobin values below the regulatory threshold).

**Methods:** The following were compared in 21,840 blood donors (aged ≥18 years) recruited from 10 mobile centres of NHSBT in England, with each method compared with the Sysmex XN-2000 haematology analyser, the reference standard: 1) gravimetry/venous HemoCue®; 2) “post donation” approach, i.e., estimating current haemoglobin concentration from that measured by a haematology analyser at a donor’s most recent prior donation; 3) capillary HemoCue®; and 4) non-invasive spectrometry (MBR Haemospect® or Orsense NMB200®). We assessed each method for sensitivity; specificity; proportion of donors who would have been inappropriately bled, or rejected from donation (“deferred”) incorrectly; and test preference.

**Results:** Compared with the reference standard, the methods ranged in test sensitivity from 17.0% (MBR Haemospect®) to 79.0% (HemoCue®) in men, and from 19.0% (MBR Haemospect®) to 82.8% (HemoCue®) in women. For specificity, the methods ranged from 87.2% (MBR Haemospect®) to 99.9% (gravimetry/venous HemoCue®) in men, and from 74.1% (Orsense NMB200®) to 99.8% (gravimetry/venous HemoCue®) in women. The proportion of donors who would have been inappropriately bled ranged from 2.2% in men for HemoCue® to 18.9% in women for MBR Haemospect®. The proportion of donors who would have been deferred incorrectly with haemoglobin concentration above the minimum threshold ranged from 0.1% in men for gravimetry/venous HemoCue® to 20.3% in women for OrSense®. Most donors preferred non-invasive spectrometry.

**Conclusion:** In the largest study reporting head-to-head comparisons of four methods to measure haemoglobin prior to blood donation, our results support replacement of venous HemoCue® with the capillary HemoCue® when donors fail gravimetry. These results have had direct translational implications for NHS Blood and Transplant in England.

## INTRODUCTION

Blood services are mandated to measure haemoglobin concentrations of potential whole blood donors in advance of each donation. The rationale is to protect the health of donors (i.e., to prevent collection from anaemic donors and mitigate the possibilities of rendering the donor anaemic) as well as to ensure the quality of blood products.^1,2^ European legislation on selection criteria of blood donors (EU directive 2004/33/EC Article 4) states that haemoglobin concentration should be ≥125 g/L for women and ≥135 g/L for men before allowing blood donation.^3^ There is, however, substantial variation across national blood services in methods of haemoglobin measurement.^4,5^ This has resulted in part because the timing of blood sampling and sample material for assessing blood donors is not defined by legislation, and partly because there is little evidence about the comparative performance of different rapid measurement methods.^6-11^

The approach of National Health Service Blood and Transplant (NHSBT, the national blood service of England) had been a gravimetric method (copper sulphate test) carried out on finger-prick capillary blood taken immediately before donation, followed by the spectrophotometric test with venous blood for those who fail the copper sulphate test.^12^ Recent data, however, indicate that this gravimetry/venous HemoCue® method may allow about 10% of donors to give blood despite having baseline haemoglobin concentrations below the minimum regulatory threshold.^13,14^ By contrast, blood services in some countries (e.g., the Netherlands and Finland) assess haemoglobin concentration before blood donation using a spectrophotometric test on capillary blood obtained by a finger-prick.^4^ Other services (e.g., France and Denmark) use haemoglobin values obtained from the most recent prior donation (“post donation” approach), employing automated haematology analysers of venous blood. ^4,15^ Other services (e.g., Bavaria, Ireland, Spain) have employed non-invasive spectrometry that does not require obtaining a blood sample.^4,16^

We conducted a within-person comparison of four haemoglobin measurement methods using performance metrics relevant to the blood donation context and comparing each method to the reference standard of a haematology analyser.

## METHODS

### Study design

This study evaluated four haemoglobin measurement approaches used by blood services in high-income countries (see “Diagnostic tests” below) against a haematology analyser reference standard. The study involved participant recruitment into two stages (**Figure 1**). Stage 1 involved direct comparisons of invasive and non-invasive methods in the same participants. Stage 2 involved an indirect comparison of two non-invasive spectrometry devices described below. Allocation of the non-invasive device between teams was done by “cross-over” randomisation. Participants in Stage 1 were not eligible to join Stage 2. The study protocol is provided in the **Annex**. The study was registered with ISRCTN (ISRCTN90871183), and approved by the National Research Ethics Service (15/EE/0335).

**Figure 1.**
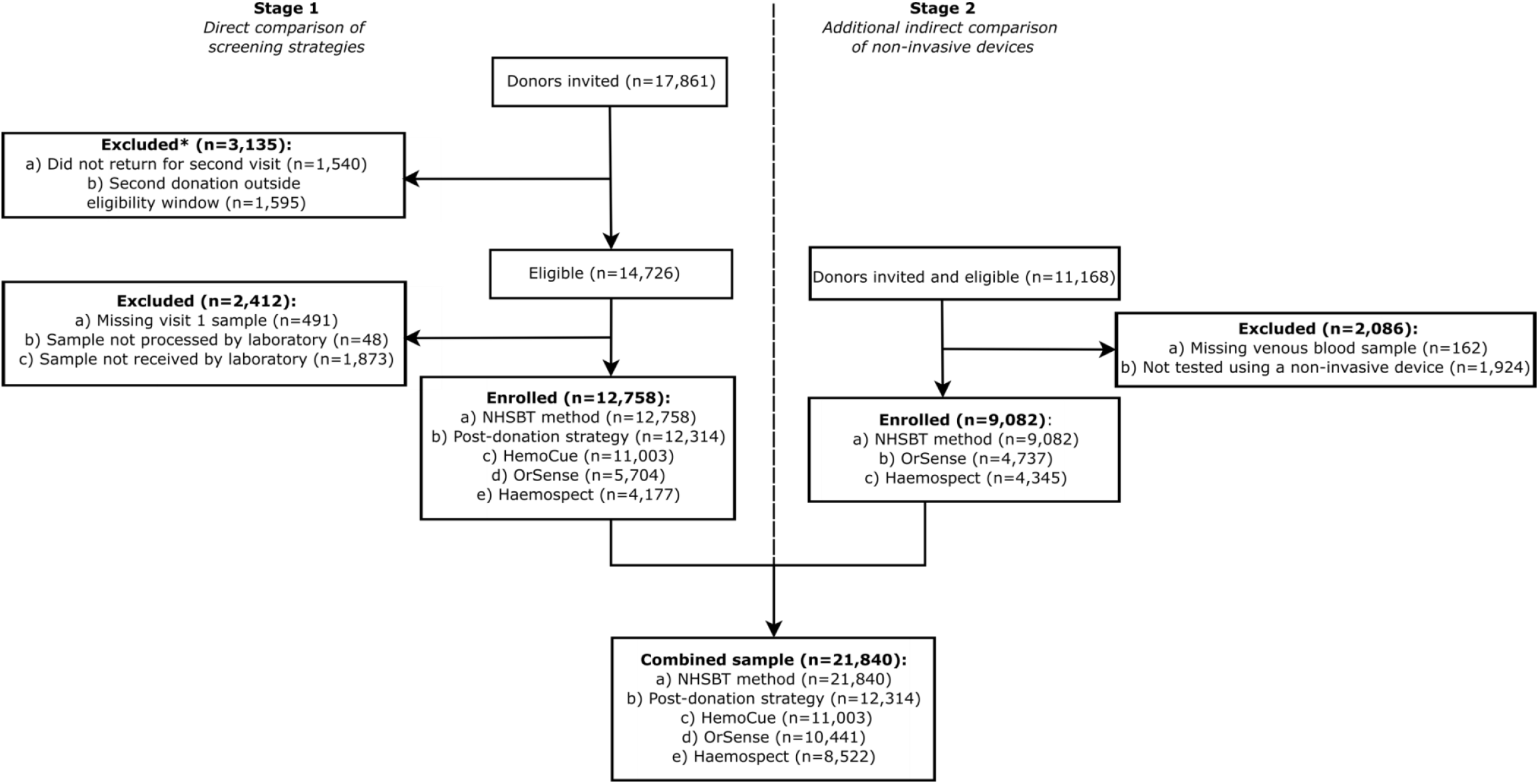
CONSORT flowchart showing. 30% drop-out rate expected between Stage 1 visit 1 and visit 2 donors as per study design.

### Diagnostic tests

We used haemoglobin concentration measured by a Sysmex XN-2000 haematology analyser at a central laboratory (UK BioCentre, Milton Keynes, UK) as the study’s reference standard.^17^ We evaluated four rapid diagnostic tests against that standard: 1) gravimetry/venous HemoCue® (NHSBT’s method at the time of this study), i.e., a copper sulphate gravimetric test carried out on finger-prick capillary blood, followed by spectrophotometry (HemoCue® AB, Ängelholm, Sweden) on venous blood for those failing gravimetry;^12^ 2) “post donation” approach, i.e., estimating current haemoglobin concentration from that measured by a haematology analyser at a donor’s most recent prior donation (i.e., about 12-16 weeks earlier); 3) a Hemocue® 301 device using finger-prick capillary blood;^18^ and 4) one of two hand-held non-invasive spectrometer devices - the MBR Haemospect® (MBR Optical Systems GmbH & Co. KG, Wuppertal, Germany)^19^ or the Orsense NMB200® (OrSense Ltd, Petah-Tikva, Israel).^20^

### Study Participants

Between February 2016 and March 2017, donors were eligible for recruitment into COMPARE if they: were aged 18 years or older; fulfilled routine criteria for donation (with the exception of pre-donation haemoglobin concentration measured using the NHSBT testing method); had an email address and access to the internet to respond to web-based questionnaires; and were willing to undergo additional haemoglobin concentration measurements at one of the 10 “mobile” donor centres of NHSBT, the sole blood provider to the NHS in England, UK. After reading study information leaflets and participating in a discussion with donor carer staff, eligible donors were asked to complete the study consent form and provide a blood sample. Soon after enrolment, participants received online health and lifestyle questionnaires, including the including the Fitzpatrick Skin Score. ^21^

### Outcomes

The primary endpoint was the proportion of donors in the study who would have been inappropriately bled by each method (i.e., the proportion of donors for whom a given method would not identify them as having sub-threshold haemoglobin levels as measured by the reference standard). Secondary endpoints included sensitivity, specificity, the proportion of donors who would have been excluded from blood donation (“deferred”) incorrectly, variability of the performance of different methods by donors’ personal characteristics (e.g., repeat versus first-time donor, and skin colour tone) and the acceptability of different methods according to donors.

### Statistical analysis

The statistical analysis followed a prespecified plan. Briefly, Bland-Altman^22^ plots were used to assess systematic difference between haemoglobin screening methods when compared against the reference standard, and supplemented by linear regression models to examine proportional biases. The percentage of donors who would have been bled below the threshold (i.e., <125 g/L for women and <135 g/L for men) was calculated by taking the number of donors categorised as having adequate haemoglobin levels by the screening method but should have been deferred according to the reference standard, and dividing by the total number of donors in the analysis population. The proportion of donors incorrectly deferred above the threshold was calculated similarly. Differences between each screening method and the reference standard were assessed using a McNemar’s test for paired within-person comparisons. For direct comparisons between strategies, donation outcomes were standardised by sex and haemoglobin level to a reference population (i.e., returning Stage 1 donor population). Each observation was assigned a weight based on the relative frequency of the sex-specific haemoglobin level appearing in the reference population relative to the estimation sample. The proportions for each of the four donation outcomes (bled below haemoglobin threshold, bled above haemoglobin threshold, deferred below haemoglobin threshold, deferred above haemoglobin threshold) were then weighted accordingly. Sensitivity (the probability of correctly identifying donors with a low haemoglobin level) and specificity (the probability of correctly identifying donors with sufficient haemoglobin levels) of each screening method was calculated and used to define the area under a receiver operating characteristic curve (AUC) to illustrate the diagnostic ability (i.e., how well a test discriminates between donors with low and sufficient haemoglobin levels) of a screening method at different haemoglobin thresholds. Sex-specific sample size was estimated to provide 80% power, at a 5% significance level, to detect a 10% relative difference in the false pass rate (i.e., percentage of donors who would have been bled below the threshold) between the NHSBT customary method and any of the other tests (**Annex**). Analyses were conducted separately for men and women using Stata v14. The analysis adhered to the Standards for Reporting Diagnostic Accuracy Studies (STARD).^23^

## RESULTS

A total of 29,029 participants were consented to participate in the COMPARE study (17,861 in Stage 1 and 11,168 in Stage 2), of whom 21,840 (75.2%) provided data for the current analysis (**Figure 1**). **Supplementary Table 1** show baseline characteristics of the participants. Compared with NHSBT’s general donor population, participants were, on average, older, more likely to be male, less ethnically diverse, and had a longer donation career (**Supplementary Tables 2** and **3**). Baseline characteristics were similar between participants recruited in Stages 1 and 2, although haemoglobin concentration was approximately 3-4 g/L lower in returning donors.

**Figure 2** and **Supplementary Figure 1** show the mean and proportional biases between the haemoglobin readings of each test and the reference standard. On average, the “post donation” approach over-estimated haemoglobin values by 3.6 g/L (limit of agreement -10.4, 17.6) and 4.0 g/L (−9.9, 18.0) for men and women, respectively, while each of the other methods tended to under-estimate haemoglobin values; -3.1 (−18.0, 11.8) and -2.4 (−16.9, 12.1) g/L for HemoCue®, -4.2 (−27.3, 18.8) and -2.2 (−30.5, 26.2) g/L for OrSense® and -6.3 (−30.7, 18.0) g/L for Haemospect®. There was evidence of proportional bias for each test, with the “post donation” approach and Haemospect® over-estimating, and HemoCue® and OrSense® underestimating haemoglobin levels at the lower end of the distribution. Mean biases for non-invasive devices were larger in donors recruited in Stage 2 (**Supplementary Figure 2**). **Figure 3** and **Supplementary Figure 3** show scatterplots of haemoglobin concentration measured by each testing method against the reference standard.

**Figure 2.**
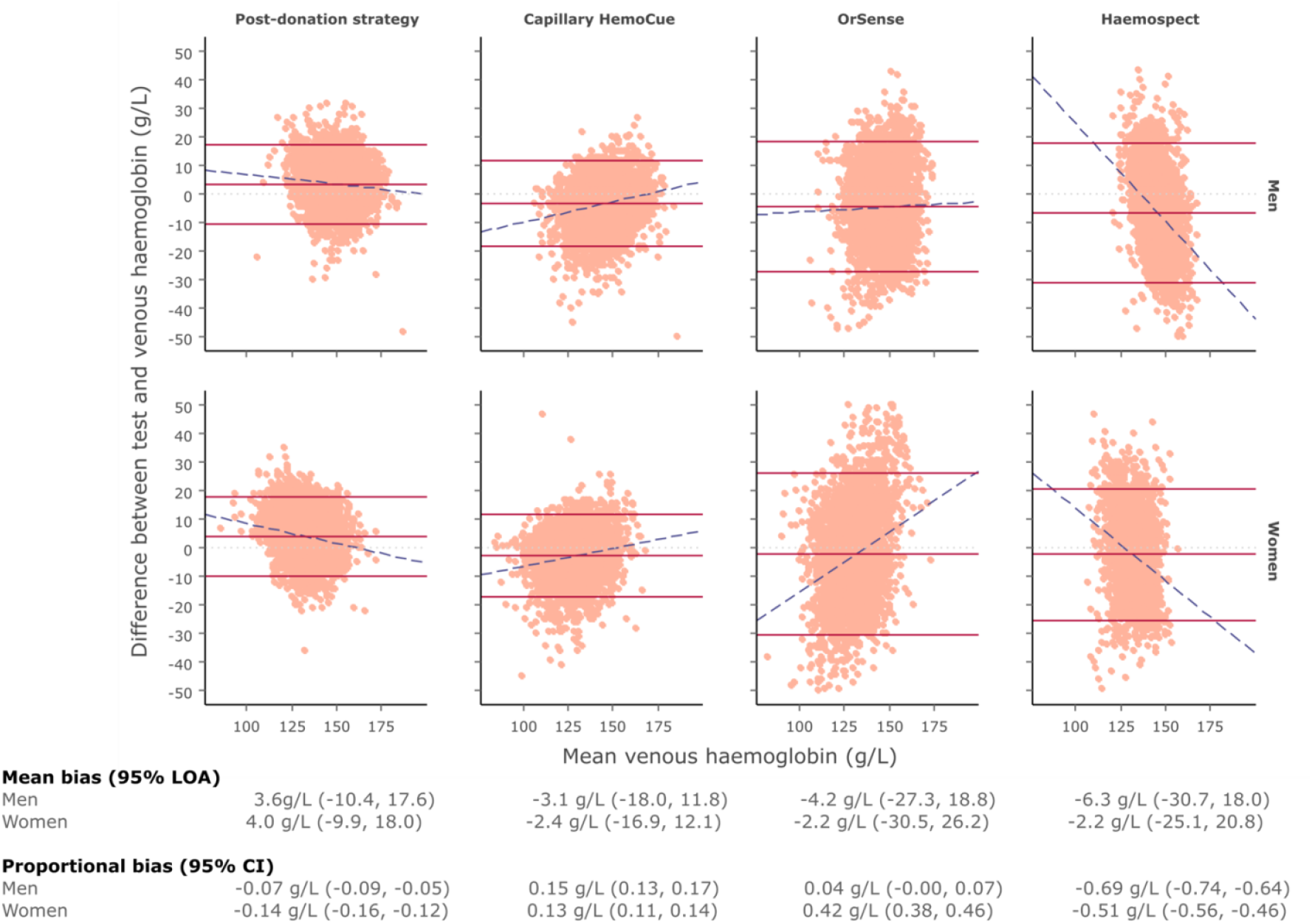
Bland-Altman plot of each haemoglobin testing strategy by sex using venous haemoglobin values as the reference test. Dotted light grey lines represents zero bias. Solid red lines represent the mean bias of the testing strategy (middle) and accompanying 95% limit of agreement (LOA; upper and lower) of the mean bias. Dashed blue lines depict proportional bias estimated using linear regression. Men - *Post-donation strategy*: N = 5920, 4.3% outside the LOA. *Capillary HemoCue*: N = 5279, 5.1% outside the LOA. *OrSense*: N = 4861, 5.7% outside the LOA. *Haemospect*: N = 4352, 5.5% outside the LOA. Women - *Post-donation strategy*: N = 6394, 5.2% outside the LOA. *Capillary HemoCue*: N = 5724, 5.0% outside the LOA. *OrSense*: N = 5580, 5.2% outside the LOA. *Haemospect*: N = 4170, 5.5% outside the LOA.

**Figure 3.**
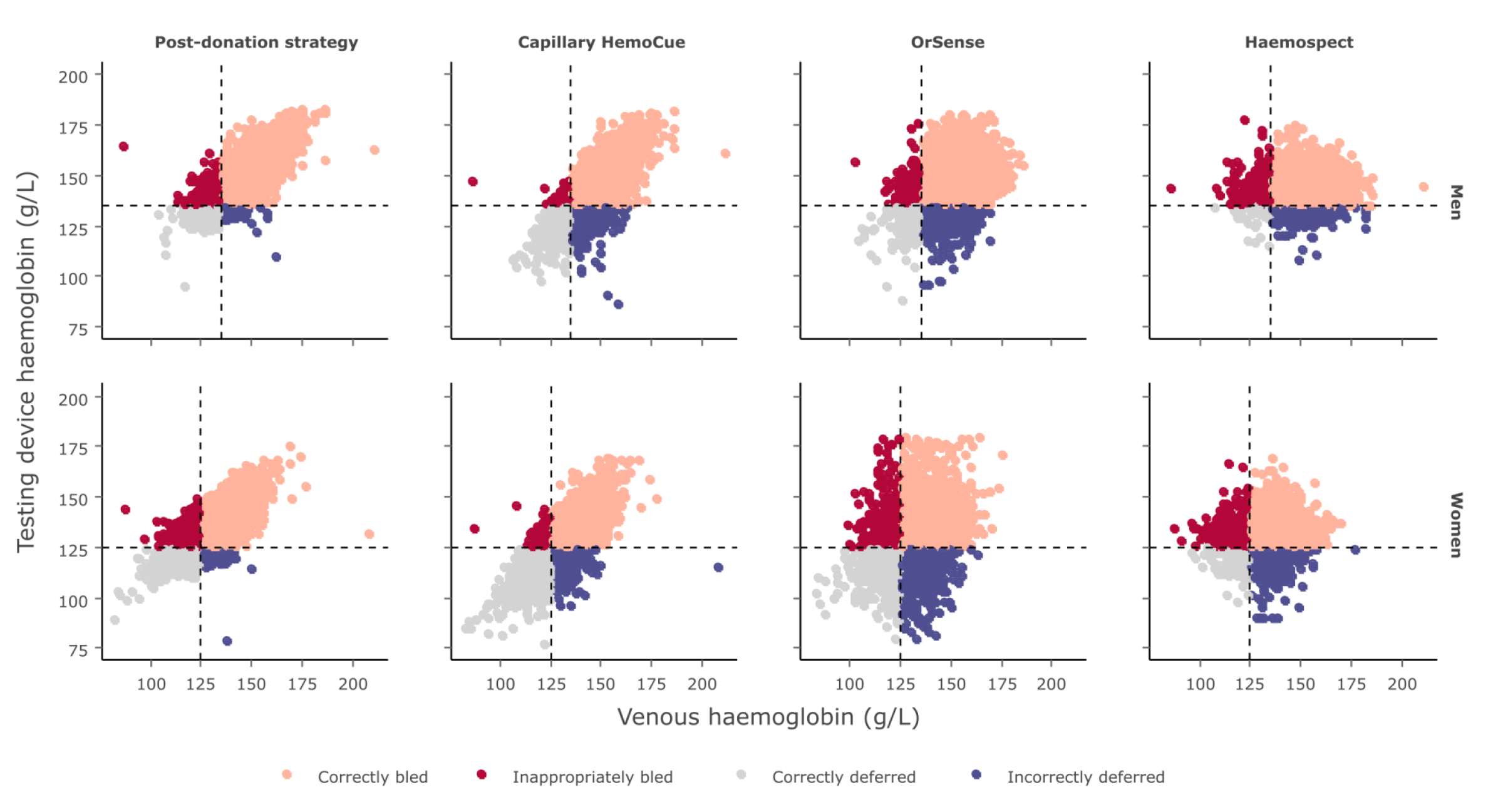
Scatterplot comparing testing device haemoglobin values to those obtained from venous blood samples by sex.

**Figure 4** shows the AUC for each test by sex. HemoCue® had the highest AUCs, for both men and women, across all haemoglobin thresholds examined, followed by the “post donation” approach, the OrSense®, and Haemospect®. The sensitivity of the different methods at minimum donation thresholds for men and women were 26.0% and 34.7% for gravimetry/venous HemoCue®, 27.9% and 35.5% for the “post donation” approach, 79.0% and 82.8% for HemoCue®, 44.4% and 51.3% for OrSense®, and 17.0% and 19.0% for Haemospect® The specificity of each method at the same haemoglobin thresholds for men and women were 99.9% and 99.8%, respectively, for gravimetry/venous HemoCue®, 98.8% and 96.6% for the “post donation” strategy, 87.6% and 82.1% for HemoCue®, 87.9% and 74.1% for OrSense®, and 87.2% for both sexes with Haemospect® (**Supplementary Figure 4**).

**Figure 4.**
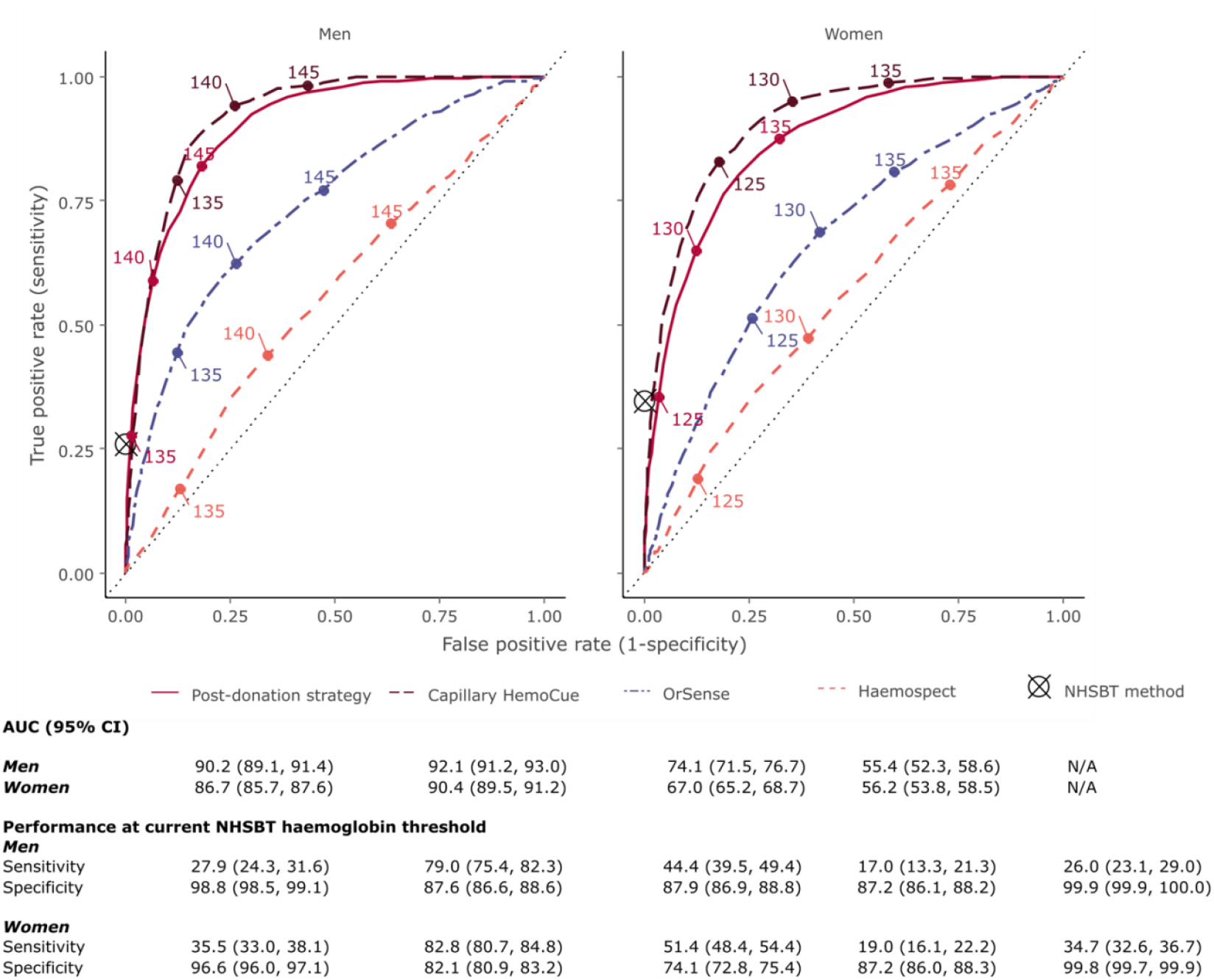
Receiver operating characteristic curves for each haemoglobin testing strategy at different haemoglobin thresholds by sex. Threshold values are shown in g/L. Sensitivity and specificity of NHSBT method has been superimposed as it only provides a pass/fail result rather than a quantitative readout.

The prevalence of donors who would have been inappropriately bled ranged from 2.2% in men for HemoCue® to 18.9% in women for MBR Haemospect® (**Figure 5** and **Supplementary Tables 4-5**). Compared to gravimetry/venous HemoCue®, use of capillary HemoCue® performed best in reducing the prevalence of inappropriate bleeding (−5·6%, -6·3, -4·9 for men and -11·1%, -11·9, -10·2 for women, p < 0.0001 for both: **Figure 6**). The proportion of donors who would have been deferred with haemoglobin concentrations above the threshold ranged from 0.1% in men for gravimetry/venous HemoCue® to 20.8% in women for OrSense® (**Figure 5** and **Supplementary Tables 4-5**). In a sensitivity analysis, the proportion of donors who would have been bled with haemoglobin concentrations below the minimum threshold using the “post donation” approach decreased whilst the number of donors inappropriately deferred somewhat increased with longer time between donation (**Supplementary Figure 5**). There were notable differences in the accuracy of methods between white and non-white donors, especially for the non-invasive devices (**Supplementary Tables 6-7** and **Supplementary Figure 6**). Stage 1 donors lost to follow-up tended to be on average younger, earlier in their donation career and more likely to have had haemoglobin values beneath the threshold at their first visit (**Supplementary Table 8**).

**Figure 5.**
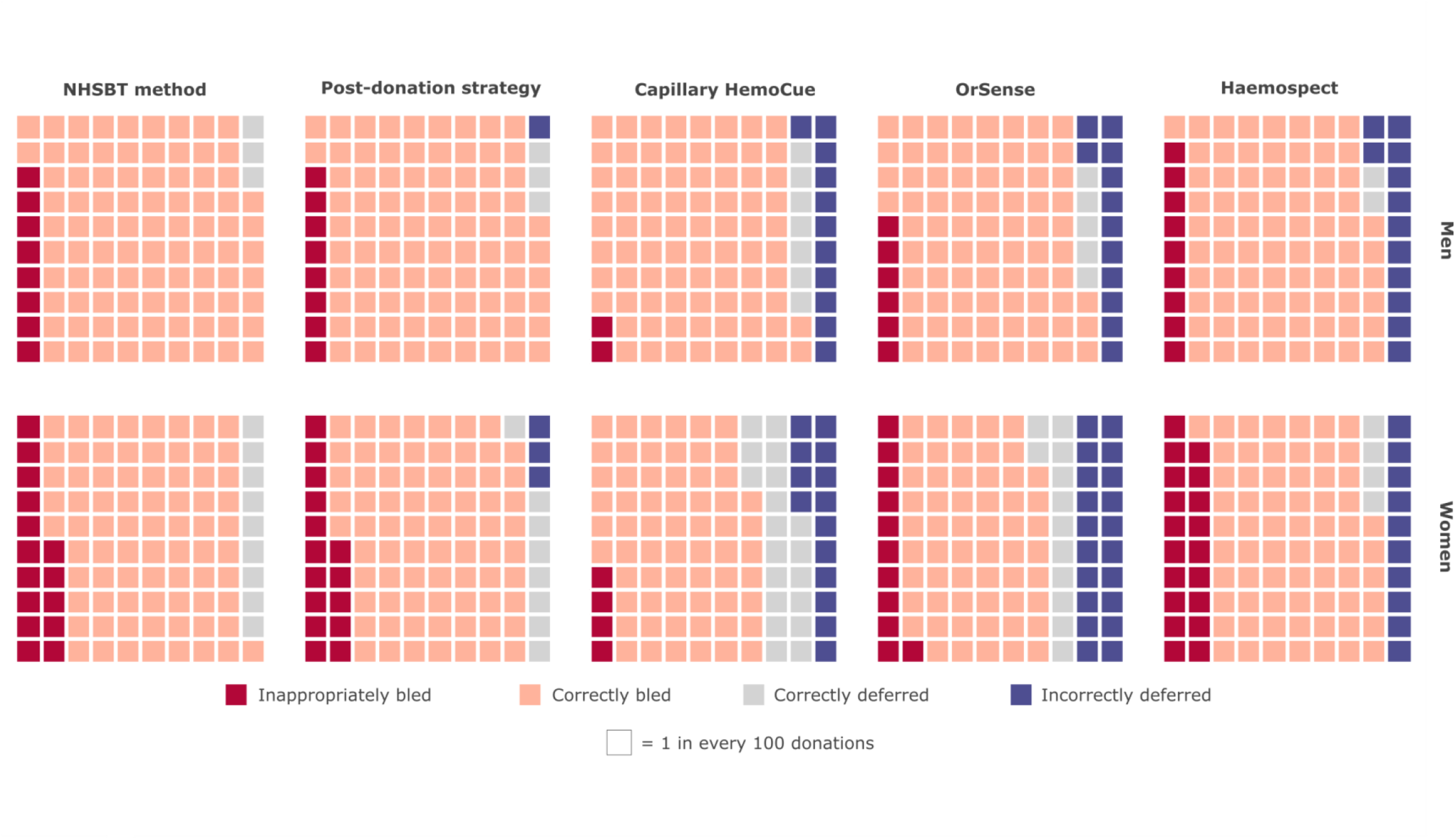
Donation outcomes by testing strategy and sex, per 100 donations standardised to the returning donor population in the COMPARE study*. * Donation outcomes by testing strategy and sex, per 100 donations standardised to the returning donor population in the COMPARE study

**Figure 6.**
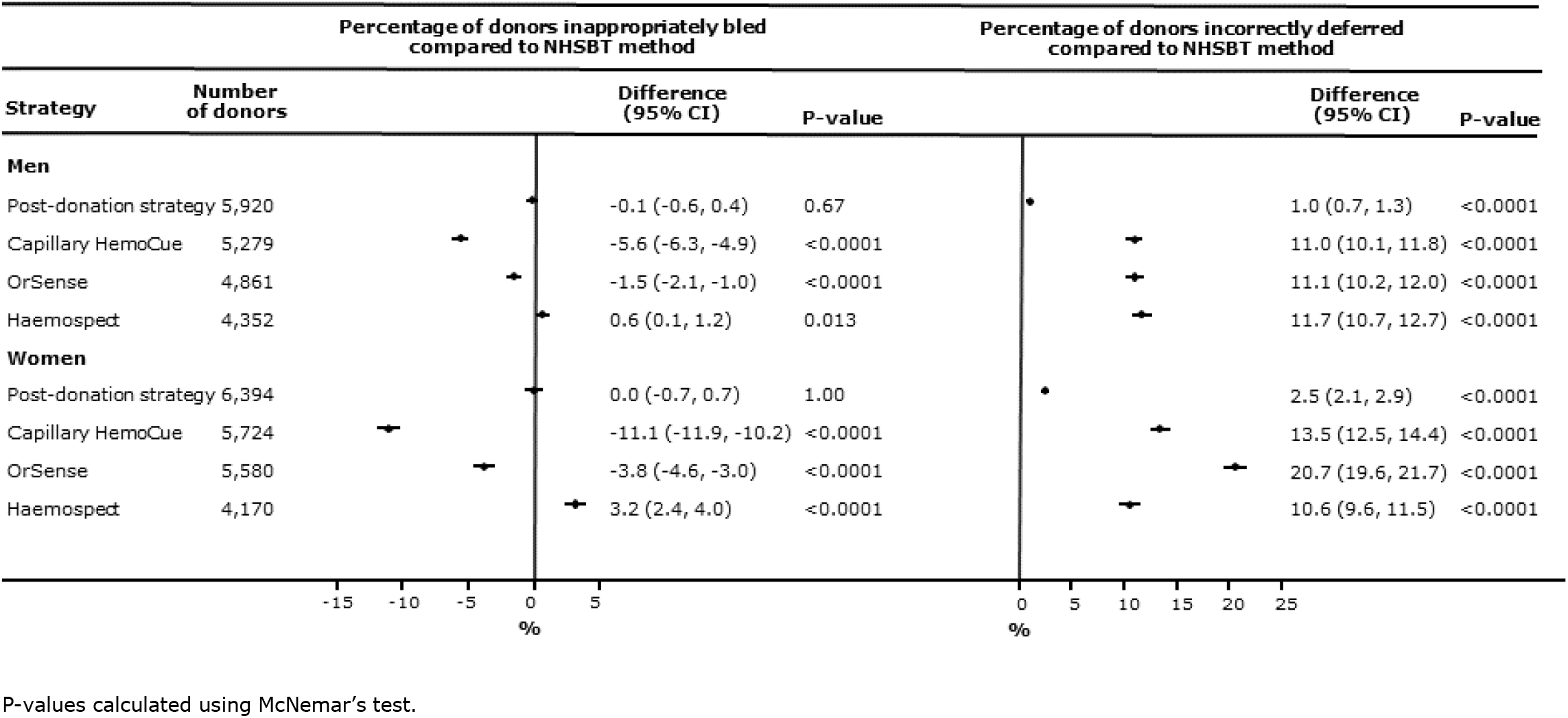
Percentage difference (95% confidence interval) in donors who would be bled below and deferred above the donation haemoglobin threshold for each testing strategy compared with the standard NHSBT test by sex. P-values calculated using McNemar’s test.

As regards test acceptability, 72% of donors preferred the non-invasive devices, 20% preferred the finger-prick test, and 8% the “post donation” approach. However, 77% of donors reported that test accuracy was their most important consideration (**Supplementary Figure 7**).

## DISCUSSION

In a study of over 21,000 whole blood donors in NHSBT, the national blood service of England, we conducted head-to-head comparisons of four rapid methods for the measurement of pre-donation haemoglobin levels, comparing each against the reference standard of a haematology analyser. We made several observations relevant to the policies and practices of blood services.

First, we found that the capillary HemoCue® method had the highest accuracy across all haemoglobin thresholds examined for both men and women, as well as the smallest biases in comparison with the reference standard. Furthermore, pre-specified subgroup analyses indicated that this method performed similarly among donors of different ages, ethnicities, and levels of blood donation experience. When compared to venous HemoCue®, use of the capillary HemoCue® method reduced the prevalence of inappropriately bled donors, but increased the proportion of donors incorrectly deferred.

When projected across the approximately 1.4 million blood donations taking place annually in England, use of the capillary HemoCue® method instead of venous HemoCue® is estimated to prevent about 65,000 donors annually from avoidable anaemia and potential iron deficiency and its potential consequences. Overall, given that safeguarding donors is the highest priority for blood services, these findings provide strong support for the replacement of venous HemoCue® with capillary HemoCue® when donors fail gravimetry, the customary method used by NHSBT. We are gratified, therefore, that in 2018 NHSBT accepted this study’s recommendation as a basis to change its haemoglobin testing practices and subsequently implemented new policies across the national blood service.^24,25^

Second, we found that the “post donation” approach (i.e., estimating current haemoglobin concentration from that measured by a haematology analyser at a donor’s most recent prior donation) performed similarly to gravimetry/venous HemoCue® when the interval between donations was about 12 to 16 weeks. However, the performance of this approach improved somewhat with longer intervals between donations, and when higher haemoglobin concentration at the first study visit were used to predict the donor’s haemoglobin concentration at the next study visit. Blood services in several countries (eg, France, Denmark and Germany) have recently adopted the “post donation” approach due to its practical advantages: it replaces the need for rapid on-site testing by using a haematology analyser at a central laboratory to measure venous blood taken from the donor’s sample pouch.^15,26,27^ Some blood services have started to supplement a post-donation approach with monitoring of serum ferritin, a measure of the body’s iron stores, in selected blood donors.^28-30^ Future work will seek to investigate the safety, cost-effectiveness, and practicability of the “post donation” approach in large, high-throughput blood services such as in England.

Third, we found that non-invasive spectrometry devices did not generally perform well in this study compared with the other methods, despite their obvious advantage of avoiding the need to take a blood sample. For example, they showed lower sensitivity for detection of haemoglobin concentration below the threshold for donation than capillary HemoCue®, meaning higher numbers of donors would be inappropriately bled. Furthermore, spectrometry devices, which measure haemoglobin by shining light on the skin of donors, performed inconsistently in people of different ethnicities and skin colour types, limiting the test’s potential applicability to blood services in countries with a large and ethnically diverse pool of donors such as in the UK. Some blood services have suffered adverse consequences from introducing non-invasive spectrometry without such robust assessment.^16^ Our study showed estimates of haemoglobin concentration by non-invasive methods would result in higher levels of inappropriate bleeding and/or higher levels of inappropriate deferral in blood donors when compared with capillary HemoCue®. Nevertheless, further efforts are warranted to improve the performance of spectrometry devices, given their potential to enhance the experience of blood donation by avoiding pain.

The current study had major strengths. It involved large numbers of participants, providing excellent statistical power and detailed comparisons of important sub-populations (e.g., sex-specific results). The study design was a within-person comparison, enhancing validity by providing head-to-head comparisons of different methods to measure haemoglobin concentrations. It involved evaluation of four methods, making it wider in scope than previous efforts focusing on fewer methods. ^7,9,31-33^ It used a state-of-the-art haematology analyser in an accredited central laboratory as the reference standard. The study was embedded in NHSBT’s routine blood service, enabling rapid recruitment of blood donors and resulting in findings of direct relevance to UK blood services.

Our study also had potential limitations. First, only about three-quarters of participants initially consented into the study returned for the second visit to allow measurements of haemoglobin concentration for the study purpose; however, a non-attendance rate of 30% at the second visit was originally factored into power calculations. Second, compared to the national donor population in England, participants in the study were older, more likely to be male, less ethnically diverse, and had a longer donation career. Hence, some caution is needed in extrapolating the findings to the general population of blood donors. Third, when assessing the post-donation approach we invited participants for a second visit about 12-16 weeks later, meaning our study had limited ability to assess this method for longer inter-donation intervals. Fourth, the study recruited only a limited number of non-white participants and relied on self-reported information for skin colour tone, limiting ability to assess potential differences by ethnic background.

In summary, in the largest study reporting head-to-head comparisons of four methods to measure haemoglobin prior to blood donation, our results support replacement of venous HemoCue® with the capillary HemoCue® when donors fail gravimetry. These results have had direct translational implications for NHS Blood and Transplant in England.

## CONTRIBUTORS

All authors contributed to data collection, study design, data analysis, interpretation, and drafting of this paper.

## Data Availability

Data are available upon request

## ACKNOWLEDGEMENTS

Participants in the COMPARE study were recruited with the active collaboration of NHS Blood and Transplant (NHSBT) England (www.nhsbt.nhs.uk). Funding was provided by NHSBT and the NIHR Blood and Transplant Research Unit (BTRU) in Donor Health and Genomics (NIHR BTRU-2014-10024). DNA extraction and genotyping were co-funded by the NIHR BTRU and the NIHR BioResource (http://bioresource.nihr.ac.uk). The academic coordinating centre for COMPARE was supported by core funding from: NIHR BTRU, UK Medical Research Council (MR/L003120/1), British Heart Foundation (RG/13/13/30194; RG/18/13/33946) and the NIHR [Cambridge Biomedical Research Centre at the Cambridge University Hospitals NHS Foundation Trust] [*]. The academic coordinating centre would like to thank blood donor centre staff and blood donors for participating in the COMPARE study.

This work was supported by Health Data Research UK, which is funded by the UK Medical Research Council, Engineering and Physical Sciences Research Council, Economic and Social Research Council, Department of Health and Social Care (England), Chief Scientist Office of the Scottish Government Health and Social Care Directorates, Health and Social Care Research and Development Division (Welsh Government), Public Health Agency (Northern Ireland), British Heart Foundation and Wellcome.

* The views expressed are those of the authors and not necessarily those of the NHS, the NIHR or the Department of Health and Social Care.

