## Supplementary Material for "Comparison of four methods to measure haemoglobin concentrations in whole blood donors (COMPARE): a diagnostic accuracy study"


| | Page |
|---|---|
| Supplementary Table 1. Participants characteristics by study stage and sex | 2 |
| Supplementary Table 2. Characteristics of NHSBT general donor population and of the COMPARE study | 3 |
| Supplementary Table 3. Differences in demographic factors of members of the COMPARE study versus the NHSBT general donor population | 4 |
| Supplementary Table 4. Standardised proportion of donors that would have been inappropriately bled or deferred using each of the testing strategies, by sex and recruitment stage | 5 |
| Supplementary Table 5. Number of donors that would have been inappropriately bled or deferred using each of the testing strategies, by sex and recruitment stage | 6 |
| Supplementary Table 6. Subgroup proportions of donors bled below the threshold for each testing strategy | 7-9 |
| Supplementary Table 7. Subgroup proportions of donors deferred above the threshold for each testing strategy | 10-12 |
| Supplementary Table 8. Baseline characteristics of analysis population in stage 1 vs those lost to follow-up | 13 |
| Supplementary Figure 1. Bland-Altman plot of each haemoglobin testing strategy using venous haemoglobin values as the reference | 14 |
| Supplementary Figure 2. Bland-Altman plot of each non-invasive haemoglobin testing strategy at stage 1 and stage 2 of the study | 15 |
| Supplementary Figure 3. Scatterplot of venous versus testing device haemoglobin values by sex | 16 |
| Supplementary Figure 4. Sensitivity and specificity of each haemoglobin testing device at haemoglobin donation threshold of 135 g/L in men and 125 g/L in women | 17 |
| Supplementary Figure 5. Forest plot of post-donation strategy performance by time taken to return for taken to return for second visit | 18 |
| Supplementary Figure 6. Non-invasive device performance in white versus non-white donors | 19 |
| Supplementary Figure 7. Donor reported preferences for each of the haemoglobin testing strategies | 20 |
| Annex. COMPARE study protocol | 21 |

**Supplementary Table 1.** Participants characteristics by study stage and sex

| | Stage 1 | | Stage 2 | |
|---|---|---|---|---|
| | **Men** | **Women** | **Men** | **Women** |
| **Number of participants[a]** | 6081 | 6677 | 4408 | 4674 |
| **Testing strategy** | | | | |
| NHSBT method | 6081 | 6677 | 4408 | 4674 |
| Post donation strategy | 5920 | 6394 | 0 | 0 |
| Capillary HemoCue | 5279 | 5724 | 0 | 0 |
| OrSense | 2646 | 3058 | 2215 | 2522 |
| Haemospect | 2159 | 2018 | 2193 | 2152 |
| **Age, years** | 50.6 (13.0) | 47.2 (13.4) | 49.6 (13.4) | 45.9 (13.7) |
| **Ethnicity[c]** | | | | |
| White | 5821 (95.9%) | 6433 (96.6%) | 4228 (96.2%) | 4558 (97.6%) |
| Mixed | 32 (0.5%) | 42 (0.6%) | 20 (0.5%) | 24 (0.5%) |
| Asian | 44 (0.7%) | 18 (0.3%) | 26 (0.6%) | 16 (0.3%) |
| Black | 13 (0.2%) | 11 (0.2%) | 15 (0.3%) | 3 (0.1%) |
| Chinese/Other | 5 (0.1%) | 6 (0.1%) | 12 (0.3%) | 9 (0.2%) |
| Prefer Not to Answer | 157 (2.6%) | 152 (2.3%) | 93 (2.1%) | 58 (1.2%) |
| **Fitzpatrick Skin Score[d]** | | | | |
| Type I (most sensitive) | 88 (2.4%) | 184 (3.9%) | 59 (2.8%) | 116 (4.7%) |
| Type II | 1300 (35.7%) | 2083 (44.5%) | 821 (39.3%) | 1154 (47.0%) |
| Type III | 2005 (55.0%) | 2206 (47.1%) | 1127 (53.9%) | 1106 (45.1%) |
| Type IV | 239 (6.6%) | 200 (4.3%) | 82 (3.9%) | 77 (3.1%) |
| Type V-VI (least sensitive) | 14 (0.4%) | 12 (0.3%) | 1 (0.1%) | 2 (0.1%) |
| **Weight, kg[b]** | 86.7 (14.1) | 73.3 (15.2) | 86.1 (14.4) | 73.6 (15.4) |
| **Blood group[e]** | | | | |
| A+ | 1873 (30.9%) | 1962 (29.5%) | 1272 (29.0%) | 1374 (29.5%) |
| A- | 492 (8.1%) | 689 (10.3%) | 370 (8.4%) | 488 (10.5%) |
| AB+ | 174 (2.9%) | 79 (1.2%) | 137 (3.1%) | 36 (0.8%) |
| AB- | 46 (0.8%) | 54 (0.8%) | 35 (0.8%) | 42 (0.9%) |
| B+ | 420 (6.9%) | 467 (7.0%) | 301 (6.9%) | 299 (6.4%) |
| B- | 129 (2.1%) | 191 (2.9%) | 110 (2.5%) | 112 (2.4%) |
| O+ | 2138 (35.2%) | 2234 (33.5%) | 1601 (36.4%) | 1621 (34.8%) |
| O- | 800 (13.2%) | 986 (14.8%) | 567 (12.9%) | 689 (14.8%) |
| **Haemoglobin concentration, g/L** | | | | |
| Baseline[f] | 150.8 (10.0) | 135.3 (9.5) | 150.7 (10.2) | 134.8 (9.5) |
| Return visit | 147.4 (10.4) | 131.4 (10.4) | - | - |
| **Haemoglobin concentration: <135g/L (M) or <125g/L (W)** | | | | |
| Baseline[f] | 286 (4.8%) | 830 (12.6%) | 237 (5.4%) | 584 (12.5%) |
| Return visit | 645 (10.6%) | 1563 (23.4%) | - | - |
| **New donor[g]** | 65 (1.1%) | 116 (1.7%) | 72 (1.6%) | 83 (1.8%) |
| **Number of blood donations in previous 2 years** | 4.1 (1.7) | 3.4 (1.6) | 3.6 (1.7) | 3.1 (1.6) |

Data presented are mean (standard deviation) or number of donors (%)

[a] Excluding 7189 donors who did not attend visit 2 (for Stage 1 donors), did not have Hb measured on the Sysmex haematology analyser (at visit 2 for Stage 1 donors, or Stage 2), or missing testing device measurement.
[b] Weight: Missing 8087 (37%)
[c] Ethnicity: Missing 44 (0.2%)
[d] The Fitzpatrick scale is a numerical classification schema for human skin colour (Fitzpatrick TB. The validity and practicality of sun-reactive skin types i through vi. Arch Dermatol 1988; 124: 869–71.) Fitzpatrick skin score (type I=Pale white skin, blue/green eyes, blond/red hair (Always burns, does not tan); type II=Fair skin, blue eyes (Burns easily, tans poorly); type III=Darker white skin (Tans after initial burn); type IV=Light brown skin (Burns minimally, tans easily); type V=Brown skin (Rarely burns, tans darkly easily); type VI=Dark brown or black skin (Never burns, always tans darkly): Missing 8964 (41%).
[e] Blood group: Missing 52 (0.2%)
[f] Baseline haemoglobin: Missing 186 (1.5%) in Stage 1 donors only
[g] A participant who has not provided a full blood donation in the five years preceding the study

**Supplementary Table 2.** Characteristics of NHSBT general donor population and of the COMPARE study*

| **Demographics**<br>n (%) or mean (SD) | **NHSBT general donor population** | **COMPARE study** |
|---|---|---|
| All donors | 949,133 (100.0) | 29,029 (100.0) |
| Sex | | |
| Male | 409,602 (43.2) | 13,650 (47.0) |
| Female | 539,531 (56.8) | 15,379 (53.0) |
| Age at baseline, years | 44.0 (15.4) | 48.4 (13.7) |
| Ethnicity | | |
| White | 875,784 (92.3) | 28,076 (96.7) |
| Asian | 25,565 (2.7) | 163 (0.6) |
| Black African | 3,261 (0.3) | 17 (0.1) |
| Black Caribbean | 3,761 (0.4) | 40 (0.1) |
| Black other | 714 (0.1) | 7 (≈0.0) |
| Chinese | 3,015 (0.3) | 21 (0.1) |
| Mixed | 12,653 (1.3) | 174 (0.6) |
| Other | 4,166 (0.4) | 28 (0.1) |
| Unknown | 20,214 (2.1) | 503 (1.7) |
| Blood group | | |
| A RhD positive (A+) | 273,533 (28.8) | 8,601 (29.6) |
| A RhD negative (A-) | 78,823 (8.3) | 2,696 (9.3) |
| AB RhD positive (AB+) | 21,880 (2.3) | 575 (2.0) |
| AB RhD negative (AB-) | 7,537 (0.8) | 253 (0.9) |
| B RhD positive (B+) | 73,594 (7.8) | 1,997 (6.9) |
| B RhD negative (B-) | 21,448 (2.3) | 722 (2.5) |
| O RhD positive (O+) | 324,592 (34.2) | 10,144 (34.9) |
| O RhD negative (O-) | 110,685 (11.7) | 4,023 (13.9) |
| Unknown | 37,041 (3.9) | 18 (0.1) |
| Donor status at baseline[a] | | |
| New | 169,429 (17.9) | 424 (1.5) |
| Occasional | 167,657 (17.7) | 3,700 (12.7) |
| More frequent | 612,047 (64.5) | 24,905 (85.8) |
| Donations (during the 2 years prior to baseline)[b] | | |
| All donations | 2.5 (2.5) | 3.5 (1.7) |
| Whole blood donations | 2.4 (2.0) | 3.5 (1.7) |
| Other donations | 0.2 (1.7) | 0.02 (0.4) |
| Deferrals (during the 2 years prior to baseline)[c] | | |
| Deferral for low haemoglobin | 56,922 (6.0) | 2,101 (7.2) |
| Any other deferral | 229,593 (24.2) | 6,999 (24.1) |
| Length of NHSBT donation history at baseline, years[d] | 10.0 (9.5) | 12.8 (9.4) |
| Venue type attended at baseline | | |
| Static centre | 173,330 (18.3) | 0 ( 0.0) |
| Mobile | 775,803 (81.7) | 29,029 (100.0) |

*NHSBT general donor 15th February 2016 to 17th March 2017

[a]"New" has been defined according to the classification used by NHSBT i.e. an individual who has not previously provided a full donation is considered to be a new donor. "Occasional" and "More frequent" have been defined as less than or equal to 2 full donations in the last 5 years and more than 2 full donations in the last 5 years, respectively.
[b] Including donations where volume equals "Normal", "Overweight" or "Missing" (not including donations where volume equals "Empty" or "Underweight").
[c] Deferrals during the 2 years prior to baseline were summarised as counts of people with a deferral for low haemoglobin levels, and more generally deferral for reasons other than low haemoglobin.
[d] Length of donation history with NHSBT at baseline was defined as the period of time between baseline and the minimum of date of registration, date of first attendance (whether successful or not) and date of first successful donation.

**Supplementary Table 3.** Differences in demographic factors of members of the COMPARE study versus the NHSBT general donor population

| **Demographics**<br>Mean or proportion (%)[a] | **COMPARE study** | **NHSBT general donor population** | **Difference (95% CI)** |
|---|---|---|---|
| Sex: Male (%) | 47.0 | 43.2 | 3.9% (3.3%, 4.4%) |
| Age at baseline, years | 48.4 | 44.0 | 4.38 (4.20, 4.56) |
| Ethnicity: White (%) | 98.4 | 94.3 | 4.1% (4.0%, 4.3%) |
| Blood group: O (%)[b] | 48.8 | 47.7 | 1.1% (0.5%, 1.7%) |
| Number of all donations[c] | 3.47 | 2.52 | 0.96 (0.93, 0.98) |
| Deferral for low haemoglobin (%)[c] | 7.2 | 6.0 | 1.2% (0.9%, 1.5%) |
| Any other deferral (%)[c] | 24.1 | 24.2 | -0.1% (-0.6%, 0.4%) |
| Length of NHSBT donation history at baseline, years | 12.8 | 9.95 | 2.81 (2.70, 2.92) |

[a] Those with missing values excluded
[b] Blood group O includes both RhD groups
[c] During the 2 years prior to baseline

**Supplementary Table 4.** Standardised proportion of donors that would have been inappropriately bled or deferred using each of the testing strategies, by sex and recruitment stage

| Testing device / strategy | Inappropriately bled | Correctly bled | Correctly deferred | Incorrectly deferred |
|---|---|---|---|---|
| **Men: Standardised** | | | | |
| NHSBT method | 7.8% | 89.3% | 2.9% | 0.1% |
| Post-donation strategy | 7.6% | 88.3% | 3.0% | 1.1% |
| Capillary HemoCue | 2.2% | 78.2% | 8.4% | 11.2% |
| OrSense | 5.8% | 77.8% | 4.8% | 11.7% |
| Haemospect | 8.7% | 77.9% | 1.9% | 11.6% |
| **Women: Standardised** | | | | |
| NHSBT method | 14.9% | 76.4% | 8.5% | 0.2% |
| Post-donation strategy | 15.0% | 73.9% | 8.4% | 2.7% |
| Capillary HemoCue | 4.1% | 62.8% | 19.3% | 13.8% |
| OrSense | 11.4% | 56.3% | 12.0% | 20.3% |
| Haemospect | 18.9% | 66.6% | 4.4% | 10.1% |

Standardised (by sex and haemoglobin level) to returning (Stage 1) donor population (N=12,758)

For direct comparisons between strategies, donation outcomes were standardised by sex and haemoglobin level to a reference population – the returning Stage 1 donor population. Each observation was assigned a weight based on the relative frequency of the sex-specific haemoglobin level appearing in the reference population relative to the estimation sample. The proportions for each of the four donation outcomes (bled below haemoglobin threshold, bled above haemoglobin threshold, deferred below haemoglobin threshold, deferred above haemoglobin threshold) were then weighted accordingly. As an example, if 200 male donors tested with Orsense had an Hb level of 120 g/l whilst only 100 male donors had this level in the reference population, each of the 200 observations would carry a weight of ½.

**Supplementary Table 5.** Number of donors that would have been inappropriately bled or deferred using each of the testing strategies, by sex and recruitment stage

| Testing device / strategy | N | Inappropriately bled | Correctly bled | Correctly deferred | Incorrectly deferred |
|---|---|---|---|---|---|
| **Men: Combined** | | | | | |
| NHSBT method | 10,489 | 653 (6.2%) | 9599 (91.5%) | 229 (2.2%) | 8 (0.1%) |
| Post-donation strategy | 5920 | 443 (7.5%) | 5243 (88.6%) | 171 (2.9%) | 63 (1.1%) |
| Capillary HemoCue | 5279 | 121 (2.3%) | 4121 (78.1%) | 455 (8.6%) | 582 (11.0%) |
| OrSense | 4861 | 224 (4.6%) | 3917 (80.6%) | 179 (3.7%) | 541 (11.1%) |
| Haemospect | 4352 | 297 (6.8%) | 3481 (80.0%) | 61 (1.4%) | 513 (11.8%) |
| **Women: Combined** | | | | | |
| NHSBT method | 11,351 | 1403 (12.4%) | 9185 (80.9%) | 744 (6.6%) | 19 (0.2%) |
| Post-donation strategy | 6394 | 926 (14.5%) | 4788 (74.9%) | 510 (8.0%) | 170 (2.7%) |
| Capillary HemoCue | 5724 | 236 (4.1%) | 3570 (62.4%) | 1137 (19.9%) | 781 (13.6%) |
| OrSense | 5580 | 535 (9.6%) | 3319 (59.5%) | 565 (10.1%) | 1161 (20.8%) |
| Haemospect | 4170 | 554 (13.3%) | 3039 (72.9%) | 130 (3.12%) | 447 (10.7%) |
| **Men: Stage 1** | | | | | |
| NHSBT method | 6081 | 467 (7.7%) | 5432 (89.3%) | 178 (2.9%) | 4 (0.1%) |
| Post-donation strategy | 5920 | 443 (7.5%) | 5243 (88.6%) | 171 (2.9%) | 63 (1.1%) |
| Capillary HemoCue | 5279 | 121 (2.3%) | 4121 (78.1%) | 455 (8.6%) | 582 (11.0%) |
| OrSense | 2646 | 170 (6.4%) | 2104 (79.5%) | 123 (4.7%) | 249 (9.4%) |
| Haemospect | 2159 | 199 (9.2%) | 1732 (80.2%) | 32 (1.5%) | 196 (9.1%) |
| **Women: Stage 1** | | | | | |
| NHSBT method | 6677 | 994 (14.9%) | 5102 (76.4%) | 569 (8.5%) | 12 (0.2%) |
| Post-donation strategy | 6394 | 926 (14.5%) | 4788 (74.9%) | 510 (8.0%) | 170 (2.7%) |
| Capillary HemoCue | 5724 | 236 (4.1%) | 3570 (62.4%) | 1137 (19.9%) | 781 (13.6%) |
| OrSense | 3058 | 394 (12.9%) | 1817 (59.4%) | 373 (12.2%) | 474 (15.5%) |
| Haemospect | 2018 | 323 (16.0%) | 1271 (63.0%) | 110 (5.5%) | 314 (15.6%) |
| **Men: Stage 2** | | | | | |
| NHSBT method | 4408 | 186 (4.2%) | 4167 (94.5%) | 51 (1.2%) | 4 (0.1%) |
| OrSense | 2215 | 54 (2.4%) | 1813 (81.9%) | 56 (2.5%) | 292 (13.2%) |
| Haemospect | 2193 | 98 (4.5%) | 1749 (79.8%) | 29 (1.3%) | 317 (14.5%) |
| **Women: Stage 2** | | | | | |
| NHSBT method | 4674 | 409 (8.8%) | 4083 (87.4%) | 175 (3.7%) | 7 (0.2%) |
| OrSense | 2522 | 141 (5.6%) | 1502 (59.6%) | 192 (7.6%) | 687 (27.2%) |
| Haemospect | 2152 | 231 (10.7%) | 1768 (82.2%) | 20 (0.9%) | 133 (6.2%) |

**Supplementary Table 6**. Subgroup proportions of donors bled below the threshold for each testing strategy

| | Men | | | Women | | |
|---|---|---|---|---|---|---|
| | **Number of donors** | **Proportion of donors bled below threshold** | **Trend p-value** | **Number of donors** | **Proportion of donors bled below threshold** | **Trend p-value** |
| **NHSBT method** | | | | | | |
| **Age group** | | | | | | |
| 18-24 | 419 (4.0%) | 4.3 (2.6, 6.7) | < 0.001 | 785 (6.9%) | 15.5 (13.1, 18.3) | < 0.001 |
| 25-29 | 548 (5.2%) | 3.8 (2.4, 5.8) | | 820 (7.2%) | 15.2 (12.9, 17.9) | |
| 30-34 | 642 (6.1%) | 3.1 (1.9, 4.8) | | 830 (7.3%) | 13.3 (11.0, 15.8) | |
| 35-39 | 634 (6.0%) | 4.4 (3.0, 6.3) | | 937 (8.3%) | 14.5 (12.3, 16.9) | |
| 40-44 | 900 (8.6%) | 5.4 (4.1, 7.1) | | 1319 (11.6%) | 13.0 (11.2, 14.9) | |
| 45-49 | 1364 (13.0%) | 4.6 (3.6, 5.9) | | 1590 (14.0%) | 13.3 (11.6, 15.0) | |
| 50-54 | 1611 (15.4%) | 6.0 (4.9, 7.3) | | 1517 (13.4%) | 11.3 (9.8, 13.0) | |
| 55-59 | 1577 (15.0%) | 6.8 (5.7, 8.2) | | 1364 (12.0%) | 9.7 (8.2, 11.4) | |
| 60-64 | 1309 (12.5%) | 7.6 (6.3, 9.2) | | 1090 (9.6%) | 11.6 (9.7, 13.6) | |
| 65-69 | 1064 (10.1%) | 10.3 (8.6, 12.3) | | 806 (7.1%) | 8.3 (6.5, 10.4) | |
| 70+ | 421 (4.0%) | 9.3 (6.7, 12.4) | | 293 (2.6%) | 10.6 (7.3, 14.7) | |
| **Ethnicity**[1] | | | | | | |
| White | 10049 (98.4%) | 6.1 (5.7, 6.6) | < 0.001 | 10991 (98.8%) | 12.3 (11.7, 12.9) | 0.007 |
| Other | 167 (1.6%) | 15.6 (10.4, 22.0) | | 129 (1.2%) | 20.2 (13.6, 28.1) | |
| **Donor status** | | | | | | |
| Returning donor | 10352 (98.7%) | 6.3 (5.8, 6.8) | 0.020 | 11152 (98.2%) | 12.4 (11.8, 13.1) | 0.099 |
| First time donor | 137 (1.3%) | 1.5 (0.2, 5.2) | | 199 (1.8%) | 8.5 (5.1, 13.3) | |
| **Number of donations In previous 2 years** | | | | | | |
| 0 | 244 (2.3%) | 2.9 (1.2, 5.8) | < 0.001 | 365 (3.2%) | 7.9 (5.4, 11.2) | 0.138 |
| 1 | 943 (9.0%) | 5 (3.7, 6.6) | | 1524 (13.4%) | 10.7 (9.2, 12.4) | |
| 2 | 1205 (11.5%) | 4.6 (3.5, 5.9) | | 1831 (16.1%) | 12.5 (11.0, 14.1) | |
| 3 | 1628 (15.5%) | 5.5 (4.5, 6.8) | | 2162 (19.0%) | 14.5 (13.1, 16.1) | |
| 4 | 2074 (19.8%) | 6.7 (5.7, 7.9) | | 2413 (21.3%) | 12.6 (11.3, 14.0) | |
| 5+ | 4395 (41.9%) | 7.2 (6.4, 8.0) | | 3056 (26.9%) | 11.9 (10.8, 13.1) | |
| **Post donation** | | | | | | |
| **Age group** | | | | | | |
| 18-24 | 233 (3.9%) | 5.6 (3.0, 9.4) | < 0.001 | 412 (6.4%) | 16.5 (13.1, 20.4) | < 0.001 |
| 25-29 | 263 (4.4%) | 5.7 (3.2, 9.2) | | 425 (6.6%) | 18.1 (14.6, 22.1) | |
| 30-34 | 340 (5.7%) | 3.2 (1.6, 5.7) | | 446 (7.0%) | 14.3 (11.2, 18.0) | |
| 35-39 | 339 (5.7%) | 5.3 (3.2, 8.3) | | 492 (7.7%) | 19.5 (16.1, 23.3) | |
| 40-44 | 505 (8.5%) | 6.9 (4.9, 9.5) | | 755 (11.8%) | 17.0 (14.3, 19.8) | |
| 45-49 | 785 (13.3%) | 5.9 (4.3, 7.7) | | 888 (13.9%) | 16.8 (14.4, 19.4) | |
| 50-54 | 942 (15.9%) | 6.9 (5.4, 8.7) | | 863 (13.5%) | 11.1 (9.1, 13.4) | |
| 55-59 | 915 (15.5%) | 7.8 (6.1, 9.7) | | 800 (12.5%) | 10.8 (8.7, 13.1) | |
| 60-64 | 736 (12.4%) | 9.8 (7.7, 12.2) | | 661 (10.3%) | 14.1 (11.5, 17.0) | |
| 65-69 | 634 (10.7%) | 11.5 (9.1, 14.3) | | 488 (7.6%) | 9.4 (7, 12.4) | |
| 70+ | 228 (3.9%) | 10.5 (6.9, 15.3) | | 164 (2.6%) | 14 (9.1, 20.3) | |
| **Ethnicity**[2] | | | | | | |
| White | 5665 (98.4%) | 7.4 (6.8, 8.1) | < 0.001 | 6160 (98.8%) | 14.5 (13.6, 15.4) | 0.751 |
| Other | 93 (1.6%) | 17.2 (10.2, 26.4) | | 76 (1.2%) | 15.8 (8.4, 26.0) | |
| **Donor status** | | | | | | |

| | | | | | | |
|---|---|---|---|---|---|---|
| Returning donor | 5855 (98.9%) | 7.5 (6.9, 8.2) | 0.175 | 6281 (98.2%) | 14.5 (13.7, 15.4) | 0.524 |
| First time donor | 65 (1.1%) | 3.1 (0.4, 10.7) | | 113 (1.8%) | 12.4 (6.9, 19.9) | |
| **Number of donations In previous 2 years** | | | | | | |
| 0 | 118 (2.0%) | 6.8 (3.0, 12.9) | 0.722 | 196 (3.1%) | 13.3 (8.9, 18.8) | < 0.001 |
| 1 | 428 (7.2%) | 5.8 (3.8, 8.5) | | 723 (11.3%) | 17.7 (15.0, 20.7) | |
| 2 | 586 (9.9%) | 7.7 (5.7, 10.1) | | 940 (14.7%) | 16.8 (14.5, 19.4) | |
| 3 | 812 (13.7%) | 8.5 (6.7, 10.6) | | 1160 (18.1%) | 16.4 (14.3, 18.6) | |
| 4 | 1199 (20.3%) | 7.7 (6.2, 9.3) | | 1413 (22.1%) | 14.2 (12.4, 16.1) | |
| 5+ | 2777 (46.9%) | 7.3 (6.4, 8.4) | | 1962 (30.7%) | 11.4 (10.0, 12.9) | |
| **Capillary HemoCue** | | | | | | |
| **Age group** | | | | | | |
| 18-24 | 203 (3.8%) | 2.5 (0.8, 5.7) | 0.039 | 365 (6.4%) | 4.7 (2.7, 7.4) | 0.165 |
| 25-29 | 243 (4.6%) | 3.3 (1.4, 6.4) | | 386 (6.7%) | 4.7 (2.8, 7.3) | |
| 30-34 | 298 (5.6%) | 0.7 (0.1, 2.4) | | 393 (6.9%) | 4.1 (2.3, 6.5) | |
| 35-39 | 310 (5.9%) | 1.6 (0.5, 3.7) | | 432 (7.5%) | 5.6 (3.6, 8.2) | |
| 40-44 | 467 (8.8%) | 1.9 (0.9, 3.6) | | 684 (11.9%) | 4.5 (3.1, 6.4) | |
| 45-49 | 679 (12.9%) | 1.9 (1.0, 3.3) | | 805 (14.1%) | 4.0 (2.7, 5.6) | |
| 50-54 | 829 (15.7%) | 2.2 (1.3, 3.4) | | 776 (13.6%) | 3.1 (2.0, 4.6) | |
| 55-59 | 831 (15.7%) | 1.6 (0.8, 2.7) | | 709 (12.4%) | 4.2 (2.9, 6.0) | |
| 60-64 | 644 (12.2%) | 2.6 (1.5, 4.2) | | 585 (10.2%) | 4.1 (2.6, 6.0) | |
| 65-69 | 564 (10.7%) | 4.3 (2.7, 6.3) | | 439 (7.7%) | 3.0 (1.6, 5.0) | |
| 70+ | 211 (4.0%) | 3.3 (1.3, 6.7) | | 150 (2.6%) | 4.7 (1.9, 9.4) | |
| **Ethnicity[3]** | | | | | | |
| White | 5049 (98.3%) | 2.3 (1.9, 2.7) | 0.147 | 5518 (98.8%) | 4.1 (3.6, 4.7) | 0.443 |
| Other | 86 (1.7%) | 4.7 (1.3, 11.5) | | 67 (1.2%) | 6.0 (1.7, 14.6) | |
| **Donor status** | | | | | | |
| Returning donor | 5219 (98.9%) | 2.3 (1.9, 2.8) | 0.233 | 5625 (98.3%) | 4.1 (3.6, 4.6) | 0.328 |
| First time donor | 60 (1.1%) | 0.0 (0.0, 6.0) | | 99 (1.7%) | 6.1 (2.3, 12.7) | |
| **Number of donations in previous 2 years** | | | | | | |
| 0 | 108 (2.0%) | 0.9 (0.0, 5.1) | 0.056 | 169 (3.0%) | 4.1 (1.7, 8.3) | 0.762 |
| 1 | 367 (7.0%) | 1.6 (0.6, 3.5) | | 624 (10.9%) | 3.5 (2.2, 5.3) | |
| 2 | 517 (9.8%) | 1.5 (0.7, 3.0) | | 830 (14.5%) | 3.7 (2.6, 5.3) | |
| 3 | 715 (13.5%) | 2.4 (1.4, 3.8) | | 1039 (18.2%) | 5.4 (4.1, 6.9) | |
| 4 | 1072 (20.3%) | 2.1 (1.4, 3.2) | | 1276 (22.3%) | 3.4 (2.5, 4.6) | |
| 5+ | 2500 (47.4%) | 2.6 (2.0, 3.3) | | 1786 (31.2%) | 4.3 (3.4, 5.3) | |
| **OrSense** | | | | | | |
| **Age group** | | | | | | |
| 18-24 | 195 (4.0%) | 4.6 (2.1, 8.6) | 0.278 | 395 (7.1%) | 14.9 (11.6, 18.8) | < 0.001 |
| 25-29 | 261 (5.4%) | 5.0 (2.7, 8.4) | | 419 (7.5%) | 15.8 (12.4, 19.6) | |
| 30-34 | 293 (6.0%) | 2.4 (1.0, 4.9) | | 406 (7.3%) | 12.3 (9.3, 15.9) | |
| 35-39 | 299 (6.2%) | 5.0 (2.8, 8.1) | | 465 (8.3%) | 11.4 (8.7, 14.6) | |
| 40-44 | 421 (8.7%) | 5.5 (3.5, 8.1) | | 634 (11.4%) | 10.7 (8.4, 13.4) | |
| 45-49 | 639 (13.1%) | 3.1 (1.9, 4.8) | | 759 (13.6%) | 11.1 (8.9, 13.5) | |
| 50-54 | 746 (15.3%) | 5.1 (3.6, 6.9) | | 783 (14.0%) | 6.6 (5.0, 8.6) | |
| 55-59 | 707 (14.5%) | 4.4 (3.0, 6.2) | | 656 (11.8%) | 6.6 (4.8, 8.7) | |
| 60-64 | 616 (12.7%) | 5.5 (3.9, 7.6) | | 518 (9.3%) | 6.6 (4.6, 9.1) | |
| 65-69 | 503 (10.3%) | 4.2 (2.6, 6.3) | | 403 (7.2%) | 5.2 (3.3, 7.9) | |

| | | | | | | |
|---|---|---|---|---|---|---|
| 70+ | 181 (3.7%) | 7.2 (3.9, 12.0) | | 142 (2.5%) | 3.5 (1.2, 8.0) | |
| **Ethnicity[4]** | | | | | | |
| White | 4662 (98.4%) | 4.5 (3.9, 5.1) | 0.001 | 5414 (98.9%) | 9.4 (8.6, 10.2) | 0.002 |
| Other | 78 (1.6%) | 12.8 (6.3, 22.3) | | 61 (1.1%) | 21.3 (11.9, 33.7) | |
| **Donor status** | | | | | | |
| Returning donor | 4795 (98.6%) | 4.7 (4.1, 5.3) | 0.228 | 5475 (98.1%) | 9.6 (8.9, 10.4) | 0.489 |
| First time donor | 66 (1.4%) | 1.5 (0.0, 8.2) | | 105 (1.9%) | 7.6 (3.3, 14.5) | |
| **Number of donations in previous 2 years** | | | | | | |
| 0 | 118 (2.4%) | 1.7 (0.2, 6.0) | 0.032 | 197 (3.5%) | 6.6 (3.6, 11.0) | 0.448 |
| 1 | 444 (9.1%) | 3.4 (1.9, 5.5) | | 788 (14.1%) | 8.4 (6.5, 10.5) | |
| 2 | 572 (11.8%) | 3.5 (2.1, 5.3) | | 898 (16.1%) | 10.7 (8.7, 12.9) | |
| 3 | 739 (15.2%) | 4.9 (3.4, 6.7) | | 1076 (19.3%) | 10.1 (8.4, 12.1) | |
| 4 | 986 (20.3%) | 5.5 (4.1, 7.1) | | 1126 (20.2%) | 9.7 (8.0, 11.6) | |
| 5+ | 2002 (41.2%) | 4.8 (3.9, 5.9) | | 1495 (26.8%) | 9.5 (8.1, 11.1) | |
| **Haemospect** | | | | | | |
| **Age group** | | | | | | |
| 18-24 | 177 (4.1%) | 4 (1.6, 8.0) | < 0.001 | 293 (7.0%) | 11.6 (8.2, 15.8) | 0.298 |
| 25-29 | 237 (5.4%) | 3.8 (1.8, 7.1) | | 305 (7.3%) | 15.1 (11.3, 19.6) | |
| 30-34 | 280 (6.4%) | 2.5 (1.0, 5.1) | | 311 (7.5%) | 10.6 (7.4, 14.6) | |
| 35-39 | 265 (6.1%) | 3.4 (1.6, 6.3) | | 358 (8.6%) | 14.8 (11.3, 18.9) | |
| 40-44 | 375 (8.6%) | 5.1 (3.1, 7.8) | | 490 (11.8%) | 16.3 (13.2, 19.9) | |
| 45-49 | 563 (12.9%) | 5.7 (3.9, 7.9) | | 603 (14.5%) | 16.7 (13.9, 20.0) | |
| 50-54 | 662 (15.2%) | 6.2 (4.5, 8.3) | | 547 (13.1%) | 11.5 (9.0, 14.5) | |
| 55-59 | 667 (15.3%) | 7.6 (5.7, 9.9) | | 480 (11.5%) | 10.0 (7.5, 13.0) | |
| 60-64 | 521 (12.0%) | 9.0 (6.7, 11.8) | | 406 (9.7%) | 11.3 (8.4, 14.8) | |
| 65-69 | 416 (9.6%) | 12.7 (9.7, 16.3) | | 274 (6.6%) | 12.4 (8.7, 16.9) | |
| 70+ | 189 (4.3%) | 11.6 (7.4, 17.1) | | 103 (2.5%) | 15.5 (9.1, 24.0) | |
| **Ethnicity[5]** | | | | | | |
| White | 4164 (98.2%) | 6.8 (6.0, 7.6) | 0.026 | 4040 (98.7%) | 13.1 (12.1, 14.2) | 0.001 |
| Other | 75 (1.8%) | 13.3 (6.6, 23.2) | | 52 (1.3%) | 28.8 (17.1, 43.1) | |
| **Donor status** | | | | | | |
| Returning donor | 4293 (98.6%) | 6.9 (6.2, 7.7) | 0.116 | 4096 (98.2%) | 13.4 (12.4, 14.5) | 0.186 |
| First time donor | 59 (1.4%) | 1.7 (0.0, 9.1) | | 74 (1.8%) | 8.1 (3.0, 16.8) | |
| **Number of donations in previous 2 years** | | | | | | |
| 0 | 104 (2.4%) | 2.9 (0.6, 8.2) | 0.002 | 136 (3.3%) | 11.8 (6.9, 18.4) | 0.501 |
| 1 | 404 (9.3%) | 4.7 (2.9, 7.2) | | 555 (13.3%) | 12.1 (9.5, 15.1) | |
| 2 | 503 (11.6%) | 6.0 (4.1, 8.4) | | 697 (16.7%) | 14.1 (11.6, 16.9) | |
| 3 | 716 (16.5%) | 6.1 (4.5, 8.2) | | 786 (18.8%) | 16.0 (13.5, 18.8) | |
| 4 | 854 (19.6%) | 7.1 (5.5, 9.1) | | 912 (21.9%) | 13.3 (11.1, 15.6) | |
| 5+ | 1771 (40.7%) | 7.9 (6.7, 9.3) | | 1084 (26.0%) | 11.6 (9.8, 13.7) | |

Data presented are proportion (95% CI) or number of donors (%). Ethnicity (NA): [1] 2.11%, [2] 2.42%, [3] 2.39%, [4] 1.95%, [5] 2.07%

**Supplementary Table 7**. Subgroup proportions of donors deferred above the threshold for each testing strategy

| | Men | | | Women | | |
|---|---|---|---|---|---|---|
| | **Number of donors** | **Proportion of donors deferred above threshold** | **Trend p-value** | **Number of donors** | **Proportion of donors deferred above threshold** | **Trend p-value** |
| **NHSBT method** | | | | | | |
| **Age group** | | | | | | |
| 18-24 | 419 (4.0%) | 0.0 (0.0, 0.9) | 0.059 | 785 (6.9%) | 0.0 (0.0, 0.5) | 0.874 |
| 25-29 | 548 (5.2%) | 0.0 (0.0, 0.7) | | 820 (7.2%) | 0.1 (0.0, 0.7) | |
| 30-34 | 642 (6.1%) | 0.0 (0.0, 0.6) | | 830 (7.3%) | 0.2 (0.0, 0.9) | |
| 35-39 | 634 (6.0%) | 0.0 (0.0, 0.6) | | 937 (8.3%) | 0.5 (0.2, 1.2) | |
| 40-44 | 900 (8.6%) | 0.0 (0.0, 0.4) | | 1319 (11.6%) | 0.1 (0.0, 0.4) | |
| 45-49 | 1364 (13.0%) | 0.1 (0.0, 0.4) | | 1590 (14.0%) | 0.1 (0.0, 0.3) | |
| 50-54 | 1611 (15.4%) | 0.1 (0.0, 0.3) | | 1517 (13.4%) | 0.2 (0.0, 0.6) | |
| 55-59 | 1577 (15.0%) | 0.1 (0.0, 0.4) | | 1364 (12.0%) | 0.2 (0.0, 0.6) | |
| 60-64 | 1309 (12.5%) | 0.3 (0.1, 0.8) | | 1090 (9.6%) | 0.3 (0.1, 0.8) | |
| 65-69 | 1064 (10.1%) | 0.1 (0.0, 0.5) | | 806 (7.1%) | 0.0 (0.0, 0.5) | |
| 70+ | 421 (4.0%) | 0.0 (0.0, 0.9) | | 293 (2.6%) | 0.0 (0.0, 1.3) | |
| **Ethnicity**[1] | | | | | | |
| White | 10049 (98.4%) | 0.1 (0.0, 0.2) | 0.716 | 10991 (98.8%) | 0.2 (0.1, 0.3) | 0.646 |
| Other | 167 (1.6%) | 0.0 (0.0, 2.2) | | 129 (1.2%) | 0.0 (0.0, 2.8) | |
| **Donor status** | | | | | | |
| Returning donor | 10352 (98.7%) | 0.1 (0.0, 0.2) | 0.745 | 11152 (98.2%) | 0.2 (0.1, 0.3) | 0.560 |
| First time donor | 137 (1.3%) | 0.0 (0.0, 2.7) | | 199 (1.8%) | 0.0 (0.0, 1.8) | |
| **Number of donations in previous 2 years** | | | | | | |
| 0 | 244 (2.3%) | 0.0 (0.0, 1.5) | 0.063 | 365 (3.2%) | 0.0 (0.0, 1.0) | 0.911 |
| 1 | 943 (9.0%) | 0.0 (0.0, 0.4) | | 1524 (13.4%) | 0.1 (0.0, 0.4) | |
| 2 | 1205 (11.5%) | 0.0 (0.0, 0.3) | | 1831 (16.1%) | 0.3 (0.1, 0.7) | |
| 3 | 1628 (15.5%) | 0.0 (0.0, 0.2) | | 2162 (19.0%) | 0.1 (0.0, 0.4) | |
| 4 | 2074 (19.8%) | 0.1 (0.0, 0.4) | | 2413 (21.3%) | 0.2 (0.1, 0.5) | |
| 5+ | 4395 (41.9%) | 0.1 (0.0, 0.3) | | 3056 (26.9%) | 0.1 (0.0, 0.3) | |
| **Post donation** | | | | | | |
| **Age group** | | | | | | |
| 18-24 | 233 (3.9%) | 1.3 (0.3, 3.7) | 0.127 | 412 (6.4%) | 1.7 (0.7, 3.5) | 0.501 |
| 25-29 | 263 (4.4%) | 0.8 (0.1, 2.7) | | 425 (6.6%) | 2.6 (1.3, 4.6) | |
| 30-34 | 340 (5.7%) | 0.3 (0.0, 1.6) | | 446 (7.0%) | 1.3 (0.5, 2.9) | |
| 35-39 | 339 (5.7%) | 0.6 (0.1, 2.1) | | 492 (7.7%) | 4.5 (2.8, 6.7) | |
| 40-44 | 505 (8.5%) | 0.4 (0.0, 1.4) | | 755 (11.8%) | 4.0 (2.7, 5.6) | |
| 45-49 | 785 (13.3%) | 0.9 (0.4, 1.8) | | 888 (13.9%) | 2.9 (1.9, 4.3) | |
| 50-54 | 942 (15.9%) | 1.6 (0.9, 2.6) | | 863 (13.5%) | 2.7 (1.7, 4.0) | |
| 55-59 | 915 (15.5%) | 1.2 (0.6, 2.1) | | 800 (12.5%) | 1.9 (1.1, 3.1) | |
| 60-64 | 736 (12.4%) | 1.6 (0.8, 2.8) | | 661 (10.3%) | 2.6 (1.5, 4.1) | |
| 65-69 | 634 (10.7%) | 0.8 (0.3, 1.8) | | 488 (7.6%) | 2.3 (1.1, 4.0) | |
| 70+ | 228 (3.9%) | 1.3 (0.3, 3.8) | | 164 (2.6%) | 1.2 (0.1, 4.3) | |
| **Ethnicity**[2] | | | | | | |
| White | 5665 (98.4%) | 1 (.7, 1.3) | 0.002 | 6160 (98.8%) | 2.7 (2.3, 3.1) | 0.497 |
| Other | 93 (1.6%) | 4.3 (1.2, 10.6) | | 76 (1.2%) | 3.9 (.8, 11.1) | |

| | | | | | | |
|---|---|---|---|---|---|---|
| **Donor status** | | | | | | |
| Returning donor | 5855 (98.9%) | 1.0 (0.8, 1.3) | 0.112 | 6281 (98.2%) | 2.7 (2.3, 3.1) | 0.237 |
| First time donor | 65 (1.1%) | 3.1 (0.4, 10.7) | | 113 (1.8%) | 0.9 (0.0, 4.8) | |
| **Number of donations in previous 2 years** | | | | | | |
| 0 | 118 (2.0%) | 3.4 (0.9, 8.5) | 0.482 | 196 (3.1%) | 1.5 (0.3, 4.4) | 0.001 |
| 1 | 428 (7.2%) | 0.7 (0.1, 2.0) | | 723 (11.3%) | 1.2 (0.6, 2.3) | |
| 2 | 586 (9.9%) | 1.5 (0.7, 2.9) | | 940 (14.7%) | 2.8 (1.8, 4.0) | |
| 3 | 812 (13.7%) | 0.9 (0.3, 1.8) | | 1160 (18.1%) | 1.8 (1.1, 2.8) | |
| 4 | 1199 (20.3%) | 0.7 (0.3, 1.3) | | 1413 (22.1%) | 2.9 (2.1, 3.9) | |
| 5+ | 2777 (46.9%) | 1.2 (0.8, 1.6) | | 1962 (30.7%) | 3.6 (2.8, 4.5) | |
| **Capillary HemoCue** | | | | | | |
| **Age group** | | | | | | |
| 18-24 | 203 (3.8%) | 7.4 (4.2, 11.9) | 0.025 | 365 (6.4%) | 13.4 (10.1, 17.4) | 0.604 |
| 25-29 | 243 (4.6%) | 7.8 (4.8, 11.9) | | 386 (6.7%) | 14.0 (10.7, 17.9) | |
| 30-34 | 298 (5.6%) | 10.4 (7.2, 14.4) | | 393 (6.9%) | 11.2 (8.3, 14.7) | |
| 35-39 | 310 (5.9%) | 10.6 (7.4, 14.6) | | 432 (7.5%) | 18.3 (14.8, 22.3) | |
| 40-44 | 467 (8.8%) | 12.6 (9.8, 16.0) | | 684 (11.9%) | 14.5 (11.9, 17.3) | |
| 45-49 | 679 (12.9%) | 10.0 (7.9, 12.5) | | 805 (14.1%) | 12.0 (9.9, 14.5) | |
| 50-54 | 829 (15.7%) | 11.2 (9.2, 13.6) | | 776 (13.6%) | 14.0 (11.7, 16.7) | |
| 55-59 | 831 (15.7%) | 11.4 (9.3, 13.8) | | 709 (12.4%) | 13.5 (11.1, 16.3) | |
| 60-64 | 644 (12.2%) | 10.1 (7.9, 12.7) | | 585 (10.2%) | 12.5 (9.9, 15.4) | |
| 65-69 | 564 (10.7%) | 14.7 (11.9, 17.9) | | 439 (7.7%) | 14.1 (11, 17.7) | |
| 70+ | 211 (4.0%) | 10.0 (6.3, 14.8) | | 150 (2.6%) | 12.7 (7.8, 19.1) | |
| **Ethnicity[3]** | | | | | | |
| White | 5049 (98.3%) | 11.0 (10.1, 11.8) | 0.842 | 5518 (98.8%) | 13.7 (12.8, 14.6) | 0.956 |
| Other | 86 (1.7%) | 11.6 (5.7, 20.3) | | 67 (1.2%) | 13.4 (6.3, 24.0) | |
| **Donor status** | | | | | | |
| Returning donor | 5219 (98.9%) | 11.1 (10.2, 11.9) | 0.503 | 5625 (98.3%) | 13.6 (12.7, 14.6) | 0.884 |
| First time donor | 60 (1.1%) | 8.3 (2.8, 18.4) | | 99 (1.7%) | 14.1 (8.0, 22.6) | |
| **Number of donations in previous 2 years** | | | | | | |
| 0 | 108 (2.0%) | 8.3 (3.9, 15.2) | 0.372 | 169 (3.0%) | 14.8 (9.8, 21.1) | 0.778 |
| 1 | 367 (7.0%) | 10.6 (7.7, 14.2) | | 624 (10.9%) | 13.9 (11.3, 16.9) | |
| 2 | 517 (9.8%) | 10.3 (7.8, 13.2) | | 830 (14.5%) | 14.1 (11.8, 16.7) | |
| 3 | 715 (13.5%) | 11.7 (9.5, 14.3) | | 1039 (18.2%) | 12.8 (10.8, 15.0) | |
| 4 | 1072 (20.3%) | 10.4 (8.7, 12.4) | | 1276 (22.3%) | 13.6 (11.8, 15.6) | |
| 5+ | 2500 (47.4%) | 11.4 (10.2, 12.7) | | 1786 (31.2%) | 13.7 (12.2, 15.4) | |
| **OrSense** | | | | | | |
| **Age group** | | | | | | |
| 18-24 | 195 (4.0%) | 4.1 (1.8, 7.9) | < 0.001 | 395 (7.1%) | 10.9 (8.0, 14.4) | < 0.001 |
| 25-29 | 261 (5.4%) | 1.9 (0.6, 4.4) | | 419 (7.5%) | 12.4 (9.4, 16.0) | |
| 30-34 | 293 (6.0%) | 5.1 (2.9, 8.3) | | 406 (7.3%) | 16.7 (13.2, 20.7) | |
| 35-39 | 299 (6.2%) | 5.4 (3.1, 8.5) | | 465 (8.3%) | 18.5 (15.1, 22.3) | |
| 40-44 | 421 (8.7%) | 10.0 (7.3, 13.2) | | 634 (11.4%) | 17.8 (14.9, 21.0) | |
| 45-49 | 639 (13.1%) | 9.5 (7.4, 12.1) | | 759 (13.6%) | 20.9 (18.1, 24.0) | |
| 50-54 | 746 (15.3%) | 10.1 (8.0, 12.4) | | 783 (14.0%) | 25.0 (22.0, 28.2) | |
| 55-59 | 707 (14.5%) | 14 (11.5, 16.8) | | 656 (11.8%) | 25.5 (22.2, 29.0) | |
| 60-64 | 616 (12.7%) | 15.4 (12.7, 18.5) | | 518 (9.3%) | 24.9 (21.2, 28.9) | |

| | | | | | | |
|---|---|---|---|---|---|---|
| 65-69 | 503 (10.3%) | 18.1 (14.8, 21.7) | | 403 (7.2%) | 25.6 (21.4, 30.1) | |
| 70+ | 181 (3.7%) | 18.8 (13.4, 25.2) | | 142 (2.5%) | 31.7 (24.1, 40.0) | |
| **Ethnicity[4]** | | | | | | |
| White | 4662 (98.4%) | 11.2 (10.3, 12.2) | 0.327 | 5414 (98.9%) | 20.9 (19.8, 22.0) | 0.034 |
| Other | 78 (1.6%) | 7.7 (2.9, 16.0) | | 61 (1.1%) | 9.8 (3.7, 20.2) | |
| **Donor status** | | | | | | |
| Returning donor | 4795 (98.6%) | 11.2 (10.3, 12.1) | 0.187 | 5475 (98.1%) | 20.9 (19.9, 22.1) | 0.057 |
| First time donor | 66 (1.4%) | 6.1 (1.7, 14.8) | | 105 (1.9%) | 13.3 (7.5, 21.4) | |
| **Number of donations in previous 2 years** | | | | | | |
| 0 | 118 (2.4%) | 7.6 (3.5, 14.0) | 0.001 | 197 (3.5%) | 19.8 (14.5, 26.1) | 0.611 |
| 1 | 444 (9.1%) | 8.3 (5.9, 11.3) | | 788 (14.1%) | 20.4 (17.7, 23.4) | |
| 2 | 572 (11.8%) | 9.3 (7.0, 11.9) | | 898 (16.1%) | 18.5 (16.0, 21.2) | |
| 3 | 739 (15.2%) | 10.4 (8.3, 12.9) | | 1076 (19.3%) | 22.9 (20.4, 25.5) | |
| 4 | 986 (20.3%) | 11.5 (9.5, 13.6) | | 1126 (20.2%) | 22.3 (19.9, 24.8) | |
| 5+ | 2002 (41.2%) | 12.6 (11.2, 14.1) | | 1495 (26.8%) | 19.9 (17.9, 22.0) | |
| **Haemospect** | | | | | | |
| **Age group** | | | | | | |
| 18-24 | 177 (4.1%) | 14.1 (9.4, 20.1) | 0.208 | 293 (7.0%) | 11.6 (8.2, 15.8) | 0.022 |
| 25-29 | 237 (5.4%) | 13.1 (9.1, 18.0) | | 305 (7.3%) | 12.8 (9.3, 17.1) | |
| 30-34 | 280 (6.4%) | 12.9 (9.2, 17.4) | | 311 (7.5%) | 13.8 (10.2, 18.2) | |
| 35-39 | 265 (6.1%) | 9.1 (5.9, 13.2) | | 358 (8.6%) | 12.0 (8.8, 15.8) | |
| 40-44 | 375 (8.6%) | 13.1 (9.8, 16.9) | | 490 (11.8%) | 10.0 (7.5, 13.0) | |
| 45-49 | 563 (12.9%) | 11.5 (9.0, 14.5) | | 603 (14.5%) | 11.3 (8.9, 14.1) | |
| 50-54 | 662 (15.2%) | 13.0 (10.5, 15.8) | | 547 (13.1%) | 9.0 (6.7, 11.7) | |
| 55-59 | 667 (15.3%) | 10.5 (8.3, 13.1) | | 480 (11.5%) | 9.6 (7.1, 12.6) | |
| 60-64 | 521 (12.0%) | 12.3 (9.6, 15.4) | | 406 (9.7%) | 9.9 (7.1, 13.2) | |
| 65-69 | 416 (9.6%) | 10.6 (7.8, 13.9) | | 274 (6.6%) | 9.5 (6.3, 13.6) | |
| 70+ | 189 (4.3%) | 10.1 (6.2, 15.3) | | 103 (2.5%) | 9.7 (4.8, 17.1) | |
| **Ethnicity[5]** | | | | | | |
| White | 4164 (98.2%) | 11.7 (10.7, 12.7) | 0.657 | 4040 (98.7%) | 10.7 (9.7, 11.7) | 0.807 |
| Other | 75 (1.8%) | 13.3 (6.6, 23.2) | | 52 (1.3%) | 9.6 (3.2, 21.0) | |
| **Donor status** | | | | | | |
| Returning donor | 4293 (98.6%) | 11.8 (10.8, 12.8) | 0.671 | 4293 (98.6%) | 11.8 (10.8, 12.8) | 0.433 |
| First time donor | 105 (1.9%) | 13.3 (7.5, 21.4) | | 59 (1.4%) | 13.6 (6.0, 25) | |
| **Number of donations in previous 2 years** | | | | | | |
| 0 | 104 (2.4%) | 14.4 (8.3, 22.7) | 0.901 | 136 (3.3%) | 12.5 (7.5, 19.3) | 0.308 |
| 1 | 404 (9.3%) | 13.1 (10, 16.8) | | 555 (13.3%) | 9.4 (7.1, 12.1) | |
| 2 | 503 (11.6%) | 10.3 (7.8, 13.3) | | 697 (16.7%) | 10.6 (8.4, 13.1) | |
| 3 | 716 (16.5%) | 11.5 (9.2, 14.0) | | 786 (18.8%) | 9.3 (7.4, 11.5) | |
| 4 | 854 (19.6%) | 11.2 (9.2, 13.6) | | 912 (21.9%) | 12.1 (10, 14.4) | |
| 5+ | 1771 (40.7%) | 12.1 (10.7, 13.8) | | 1084 (26.0%) | 11.2 (9.3, 13.2) | |

Data presented are proportion (95% CI) or number of donors (%)

Ethnicity (NA): [1] 2.11%, [2] 2.42%, [3] 2.39%, [4] 1.95%, [5] 2.07%

**Supplementary Table 8**. Baseline characteristics of analysis population in stage 1 vs those lost to follow-up

| | Men | | | Women | | |
|---|---|---|---|---|---|---|
| | **Analysis** | **Lost to follow-up** | **P-value** | **Analysis** | **Lost to follow-up** | **P-value** |
| **Number of participants** | 6081 | 2120 | | 6677 | 2983 | |
| **Age, years** | 50.6 (13.0) | 48.0 (14.3) | 0.00 | 47.2 (13.4) | 44.4 (14.2) | 0.00 |
| **Weight, kg**[1] | 86.7 (14.1) | 86.0 (14.3) | 0.14 | 73.3 (15.2) | 72.9 (15.3) | 0.30 |
| **Ethnicity**[2] | | | | | | |
| White | 5821 (95.9%) | 2015 (95.9%) | | 6433 (96.6%) | 2855 (96.6%) | |
| Mixed | 32 (0.5%) | 13 (0.6%) | | 42 (0.6%) | 24 (0.8%) | |
| Asian | 44 (0.7%) | 18 (0.9%) | 0.256 | 18 (0.3%) | 20 (0.7%) | 0.002 |
| Black | 13 (0.2%) | 3 (0.1%) | | 11 (0.2%) | 9 (0.3%) | |
| Chinese/Other | 5 (0.1%) | 6 (0.3%) | | 6 (0.1%) | 5 (0.2%) | |
| Prefer Not to Answer | 147 (2.6%) | 47 (2.2%) | | 152 (2.3%) | 43 (1.5%) | |
| **Fitzpatrick Skin Score**[3] | | | | | | |
| Type I (Most sensitive) | 88 (2.4%) | 24 (2.5%) | | 59 (3.9%) | 184 (3.9%) | |
| Type II | 1300 (35.7%) | 363 (37.7%) | | 645 (42.2%) | 2083 (44.5%) | |
| Type III | 2005 (55.0%) | 515 (53.5%) | 0.798 | 753 (49.3%) | 2206 (47.1%) | 0.494 |
| Type IV | 239 (6.6%) | 58 (6.0%) | | 68 (4.5%) | 200 (4.3%) | |
| Type V-VI (Least sensitive) | 14 (0.4%) | 3 (0.3%) | | 2 (0.1%) | 12 (0.3%) | |
| **Blood group**[4] | | | | | | |
| A+ | 1873 (30.9%) | 640 (30.5%) | | 1962 (29.5%) | 872 (29.6%) | |
| A- | 492 (8.1%) | 178 (8.5%) | | 689 (10.3%) | 273 (9.3%) | |
| AB+ | 174 (2.9%) | 64 (3.1%) | | 79 (1.2%) | 48 (1.6%) | |
| AB- | 46 (0.8%) | 20 (1.0%) | 0.895 | 54 (0.8%) | 32 (1.1%) | 0.368 |
| B+ | 420 (6.9%) | 133 (6.3%) | | 467 (7.0%) | 215 (7.3%) | |
| B- | 129 (2.1%) | 43 (2.1%) | | 191 (2.9%) | 89 (3.0%) | |
| O+ | 2138 (35.2%) | 759 (36.1%) | | 2234 (33.5%) | 990 (33.6%) | |
| O- | 800 (13.2%) | 264 (12.6%) | | 986 (14.8%) | 430 (14.6%) | |
| **Haemoglobin concentration, g/L** | | | | | | |
| Baseline[5] | 150.8 (10.0) | 150.5 (10.4) | 0.236 | 135.3 (9.5) | 134.2 (10.6) | 0.000 |
| **Haemoglobin concentration: <135g/L (men) or <125g/L (women)** | | | | | | |
| Baseline[5] | 286 (4.8%) | 122 (5.9%) | 0.047 | 830 (12.6%) | 461 (15.9%) | 0.000 |
| **New donor**[6] | 65 (1.1%) | 83 (3.9%) | 0.000 | 116 (1.7%) | 125 (4.2%) | 0.000 |
| **Number of blood donations in previous 2 years** | 4.1 (1.7) | 3.5 (1.8) | 0.000 | 3.4 (1.6) | 2.9 (1.7) | 0.000 |

Data presented are mean (standard deviation) or number of donors (%)
P-value calculated using a t-test for continuous variables and a chi-squared test for categorical variables.

[1]Weight: Missing 6226 (35%)
[2]Ethnicity: Missing 69 (0.4%)
[3]Fitzpatrick score: Missing 7040 (39%)
[4]Blood group: Missing 77 (0.4%)
[5] Baseline haemoglobin: Missing 308 (1.7%) in Stage 1 donors only
[6]A participant who has not provided a full blood donation in the five years preceding the study.

**Supplementary Figure 1**. Bland-Altman plot of each haemoglobin testing strategy using venous haemoglobin values as the reference

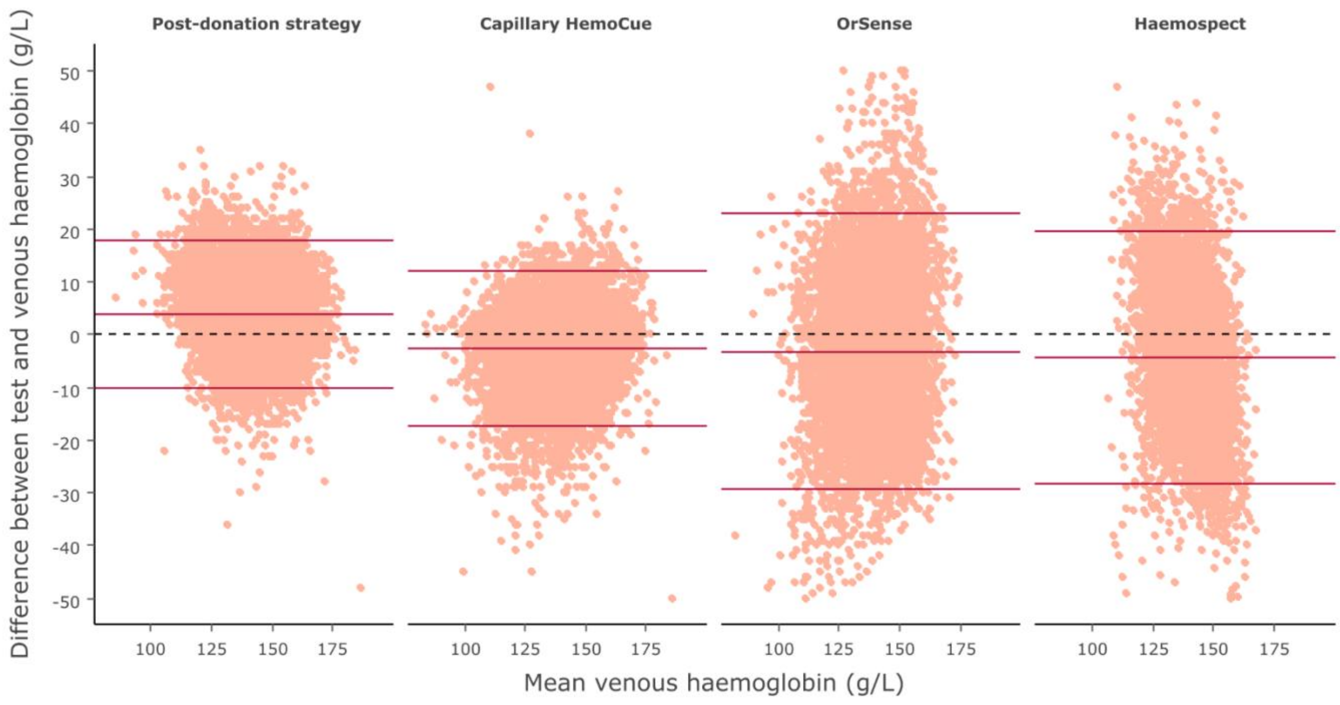


Dotted black reference line represents zero bias. Solid red lines represent the mean bias of the testing strategy (middle) and accompanying 95% limit of agreement (LOA) of the mean bias (upper and lower).

Post-donation strategy: N = 12314, mean bias = 3.8g/L (3.7, 4.0), 95% LOA = (-10.1, 17.8), 4.4% outside the LOA.
Capillary HemoCue: N = 11033, mean bias = -2.7g/L (-2.9, -2.6), 95% LOA = (-17.4, 12.0), 4.9% outside the LOA.
OrSense: N = 10441, mean bias = -3.1g/L (-3.4, -2.9), 95% LOA = (-29.2, 22.9), 5.3% outside the LOA.
Haemospect: N = 8522, mean bias = -4.3g/L (-4.6, -4.0), 95% LOA = (-28.3, 19.7), 5.4% outside the LOA.

**Supplementary Figure 2**. Bland-Altman plot of each non-invasive haemoglobin testing strategy at stage 1 and stage 2 of the study

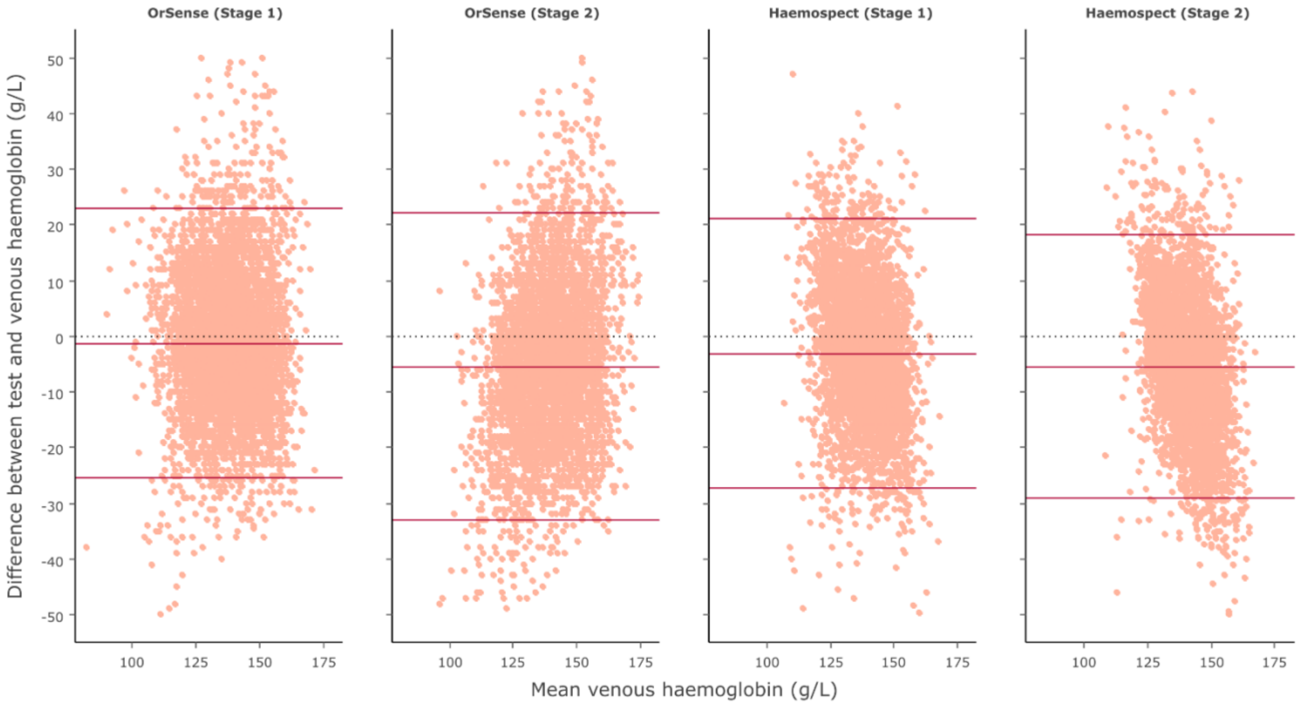


Dotted black reference line represents zero bias. Solid red lines represent the mean bias of the testing strategy (middle) and accompanying 95% limit of agreement (LOA) of the mean bias (upper and lower).

OrSense stage 1: N = 5704, mean bias = -1.2g/L (-1.6, -0.9), 95% LOA = (-25.4, 22.9), 5.3% outside the LOA.
OrSense stage 2: N = 4737, mean bias = -5.4g/L (-5.8, -5.0), 95% LOA = (-33.0, 22.1), 5.4% outside the LOA.
Haemospect stage 1: N = 4177, mean bias = -3.1g/L (-3.5, -2.7), 95% LOA = (-27.2, 21.1), 5.0% outside the LOA.
Haemospect stage 2: N = 4345, mean bias = -5.5g/L (-5.8, -5.1), 95% LOA = (-29.1, 18.2), 5.7% outside the LOA.

**Supplementary Figure 3.** Scatterplot of venous versus testing device haemoglobin values by sex

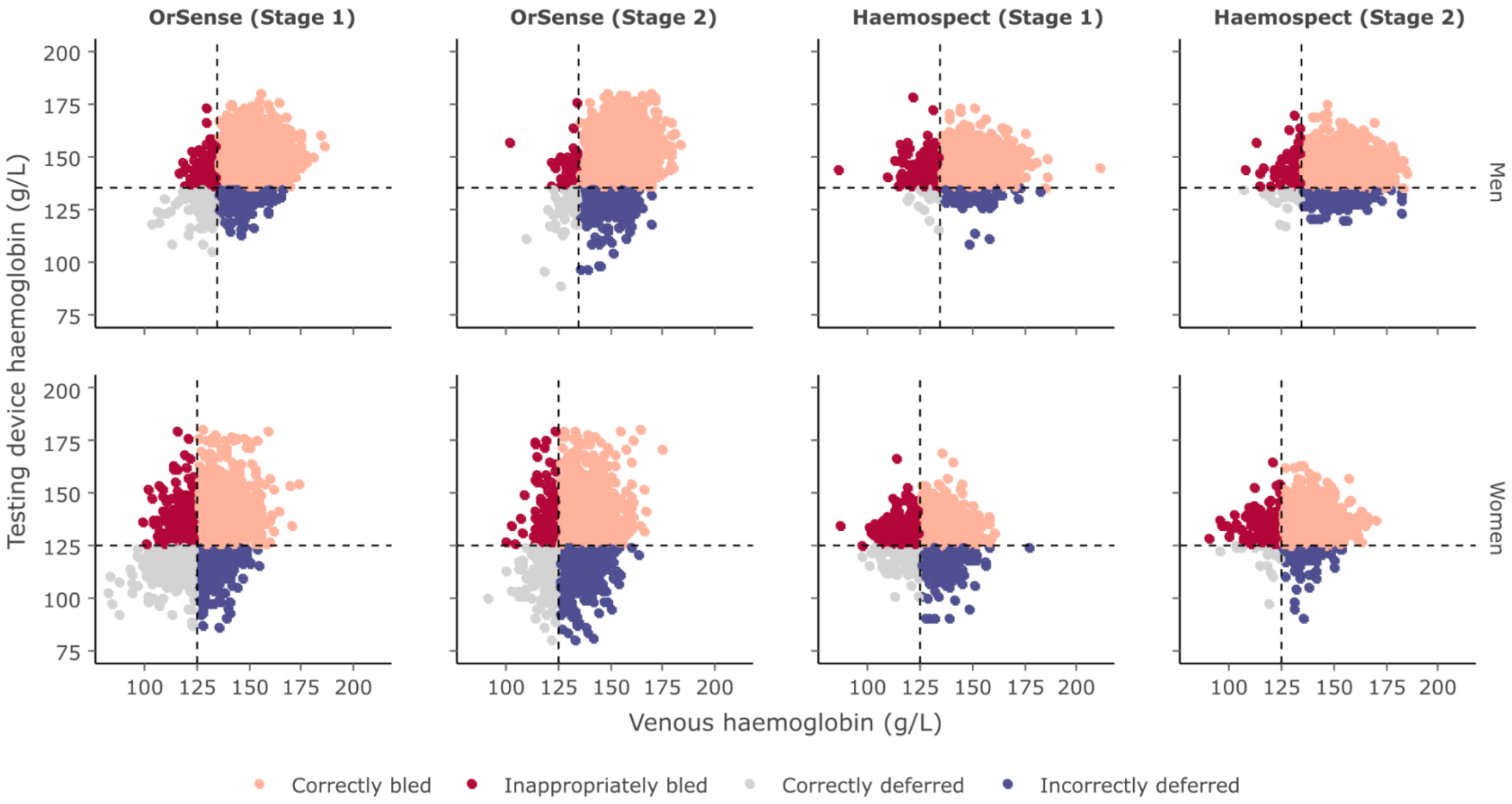

**Supplementary Figure 4**. Sensitivity and specificity of each haemoglobin testing device at haemoglobin donation threshold of 135 g/L in men and 125 g/L in women

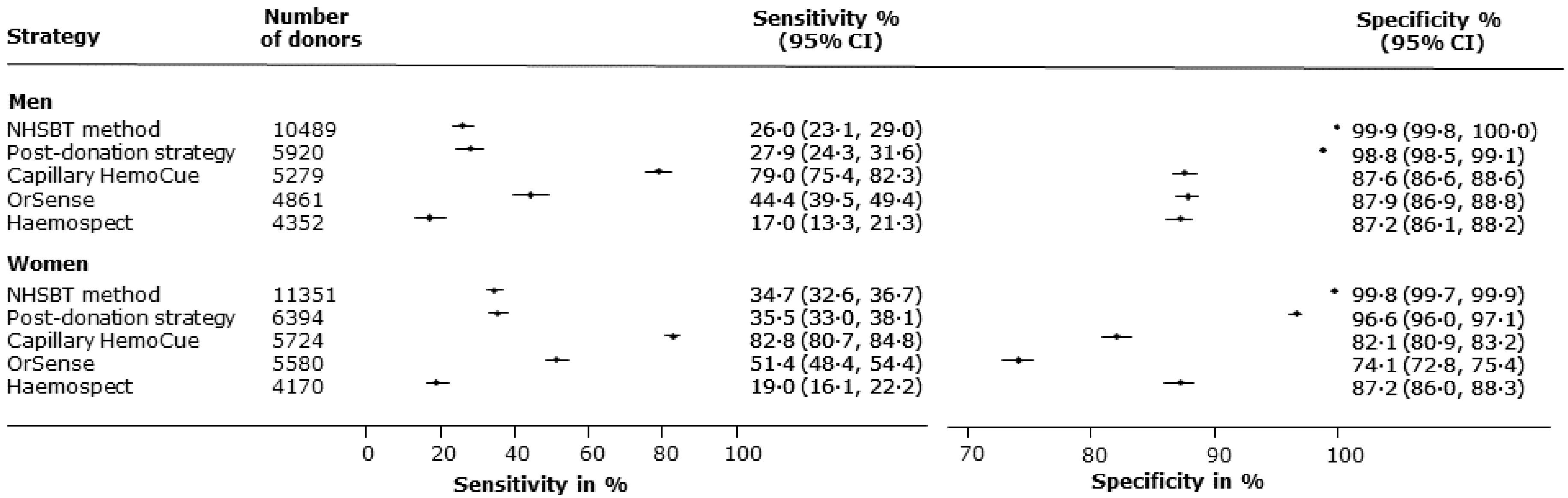

**Supplementary Figure 5**. Forest plot of post-donation strategy performance by time taken to return for taken to return for second visit

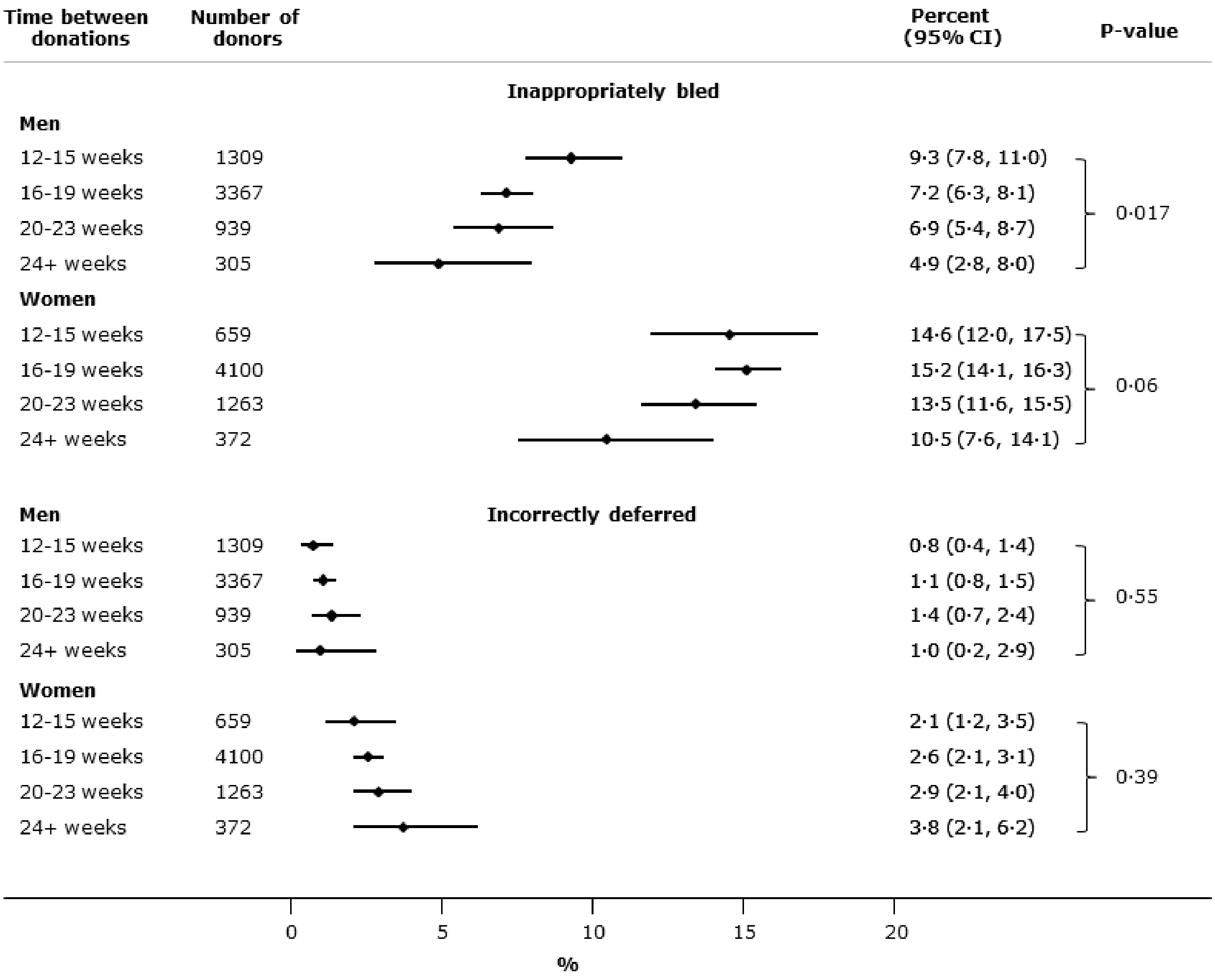

**Supplementary Figure 6**. Non-invasive device performance in white versus non-white donors

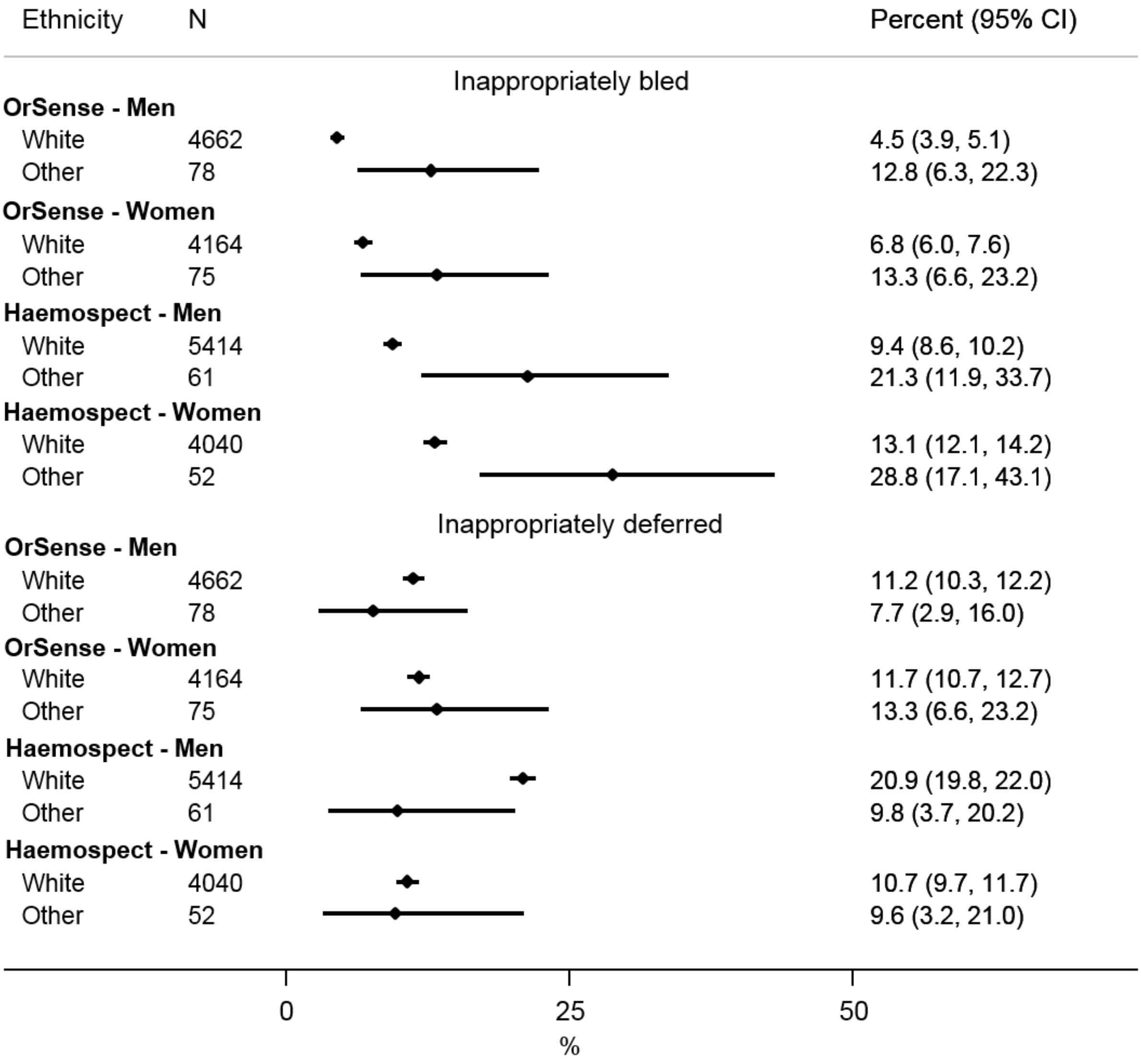

**Supplementary Figure 7**. Donor reported preferences for each of the haemoglobin testing strategies

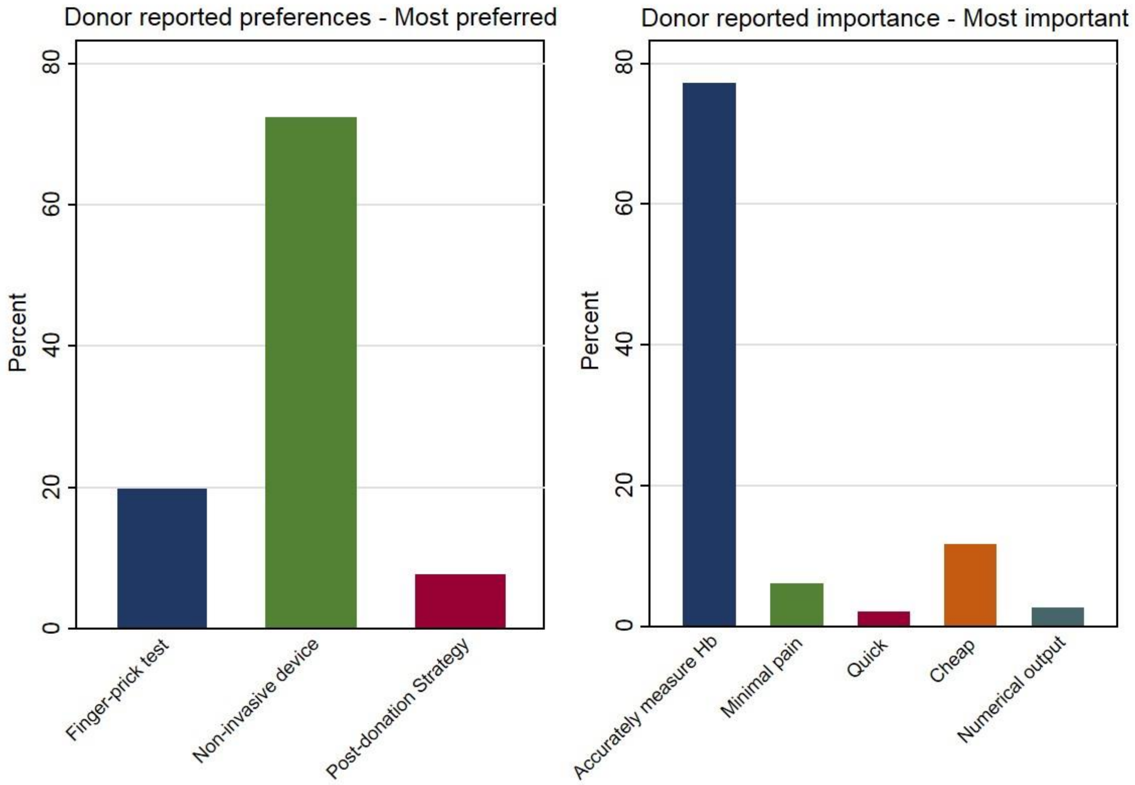
